# Quantifying the contribution of genetic variation to healthcare expenditure across diverse healthcare systems

**DOI:** 10.64898/2026.07.14.26358031

**Authors:** Sebastian May-Wilson, Jiwoo Lee, Tomoko Nakanishi, Camiel van der Laan, Ioannis Louloudis, Kuang Lin, Stavroula Kanoni, Patrick Fahr, Anne Richmond, Chadi Saad, Penelope A. Lind, Zaina Al-Kanaani, Mykyta Artomov, Karina Banasik, Enda M. Byrne, Zhengming Chen, Christian Erikstrup, Erik Sørensen, Jakob German, Giulia Brunelli, Daníel F. Guðbjartsson, Unnur Þorsteinsdóttir, Ian B. Hickie, Nikita Kolosov, Satoshi Koyama, Arne Kukkonen, Liming Li, Daniel L McCartney, Laust Hvas Mortensen, Sisse Rye Ostrowski, Ole B. Pedersen, Henning Bundgaard, Dan J. Siskind, Doug Speed, Patrick Sulem, Andres Võrk, Sarah Wordsworth, Zhiyu Yang, Kristina Zguro, Genes & Health Research Team, FinnGen, Sarah E. Medland, Nicholas G. Martin, Hamdi Mbarek, Dorret I. Boomsma, Riccardo E Marioni, James Buchanan, Pradeep Natarajan, Sarah Finer, David A van Heel, Robin Walters, Erik Abner, Søren Brunak, Neil Martin Davies, Tatiana Cajuso Pons, Padraig Dixon, Andrea Ganna

## Abstract

Healthcare systems must balance rising costs with the delivery of effective care, yet the factors underlying large inter-individual differences in healthcare expenditure remain incompletely understood. Here we examine how genome-wide genetic variation contributes to healthcare costs, analysing inpatient, outpatient, primary care and prescription drug expenditure in up to 1,429,889 individuals from 11 studies across 7 countries.

We identify hundreds of common genetic variants robustly associated with healthcare costs, revealing a reproducible polygenic architecture shared across healthcare systems. Individual common variants have modest effects, typically altering annual costs by ∼1-2% per allele, with the strongest signals arising from the HLA region, consistent with pleiotropic effects across autoimmune and inflammatory diseases. In contrast, putative loss-of-function (pLOF) variants (ClinVar/ENIGMA pathogenic variants or LOFTEE high-confidence pLOF) in clinically actionable genes, studied in UK Biobank, have large individual-level consequences: carriers of such variants in BRCA1, BRCA2, MSH2 and APC experience more than a two-fold increase in annual inpatient costs.

Cost-associated signals colocalize extensively with autoimmune disorders, cardiometabolic risk factors, pain sensitivity, and depression amongst others. Polygenic scores derived for healthcare costs can predict up to 1.4% of drug-related healthcare expenditure in independent cohorts and retain their effects in within-family analyses, indicating largely direct genetic influences. By linking genetic risk to healthcare expenditure, this work provides a foundation for integrating human genetics into health economics, preventive strategies and population-level screening.

## Introduction

Healthcare expenditure represents a major component of economic activity, averaging 9.2% of GDP across OECD member countries^1^. Understanding the determinants of healthcare costs is essential for prioritizing prevention strategies and optimizing the allocation of healthcare resources^2^.

Previous work has linked healthcare spending to disease burden. In the United States, type 2 diabetes accounts for the largest share of disease-specific spending, followed by musculoskeletal disorders and ischemic heart disease^3^. Much of this burden is linked to modifiable risk factors such as obesity, hypertension, hyperglycaemia, poor diet, and smoking^4^. Multimorbidity further amplifies costs, with co-occurring chronic conditions generating super-additive increases in healthcare expenditure^5^.

Despite these advances, the role of non-modifiable genetic risk factors in healthcare expenditure remains less explored. Genetic variation may influence healthcare costs both by predisposing individuals to specific diseases and through pleiotropic effects that affect multiple traits and conditions. Twin and family-based studies suggest that healthcare costs are partly heritable, with heritability estimates of 0.29 in the U.S.^6^ and 0.29–0.37 in the Netherlands^7^. Yet, a direct mapping between genetic variation and healthcare costs has yet to be undertaken.

However, why should genetic variants be linked to healthcare expenditure, given that genetic factors are largely non-modifiable?

First, directly estimating the contribution of genetic risk to healthcare expenditure provides an empirical foundation for economic models of healthcare costs and helps clarify when and for whom genetic information meaningfully contributes to population health. For example, quantifying expenditure associated with pathogenic variants or elevated polygenic risk can inform the cost-effectiveness of screening and other interventions. Although polygenic risk scores (PGSs) have attracted substantial interest as predictive screening tools^8–12^, their clinical benefit remains uncertain. Economic evaluations suggested PGS-informed screening or prevention strategies can appear cost-effective, but the evidence base is dominated by hypothetical cohorts and disease-specific endpoints^13,14^.

Second, longitudinal healthcare cost data provide an integrated measure of an individual’s morbidity trajectory, reflecting the cumulative effects of chronic disease, acute events, behaviours, and social factors. By identifying genetic associations with these costs, we may uncover biological or behavioural pathways driving illness, multimorbidity and persistent high healthcare use that may not otherwise be identified. This could shed light on why targeted programs for “super-utilizers” of healthcare resources have shown limited success^15^.

Third, genetic instruments can be leveraged in causal inference frameworks to estimate the causal impact of modifiable risk factors on healthcare costs, as demonstrated for adiposity, smoking and blood pressure^16–19^. However, these studies have focused on a limited set of risk factors, underscoring the need for a broader understanding of how genetic variation across a wide range of traits influences healthcare expenditure.

## Results

### Phenotype and study characteristics

We considered 11 studies participating in the GenCost consortium (**Supplementary Tables 1 and 2**). The combined dataset included up to 1,429,889 individuals for the largest phenotype from 11 studies across 7 countries. (**Figure 1A**), with seven of these cohorts originating from Europe.

**Figure 1.**
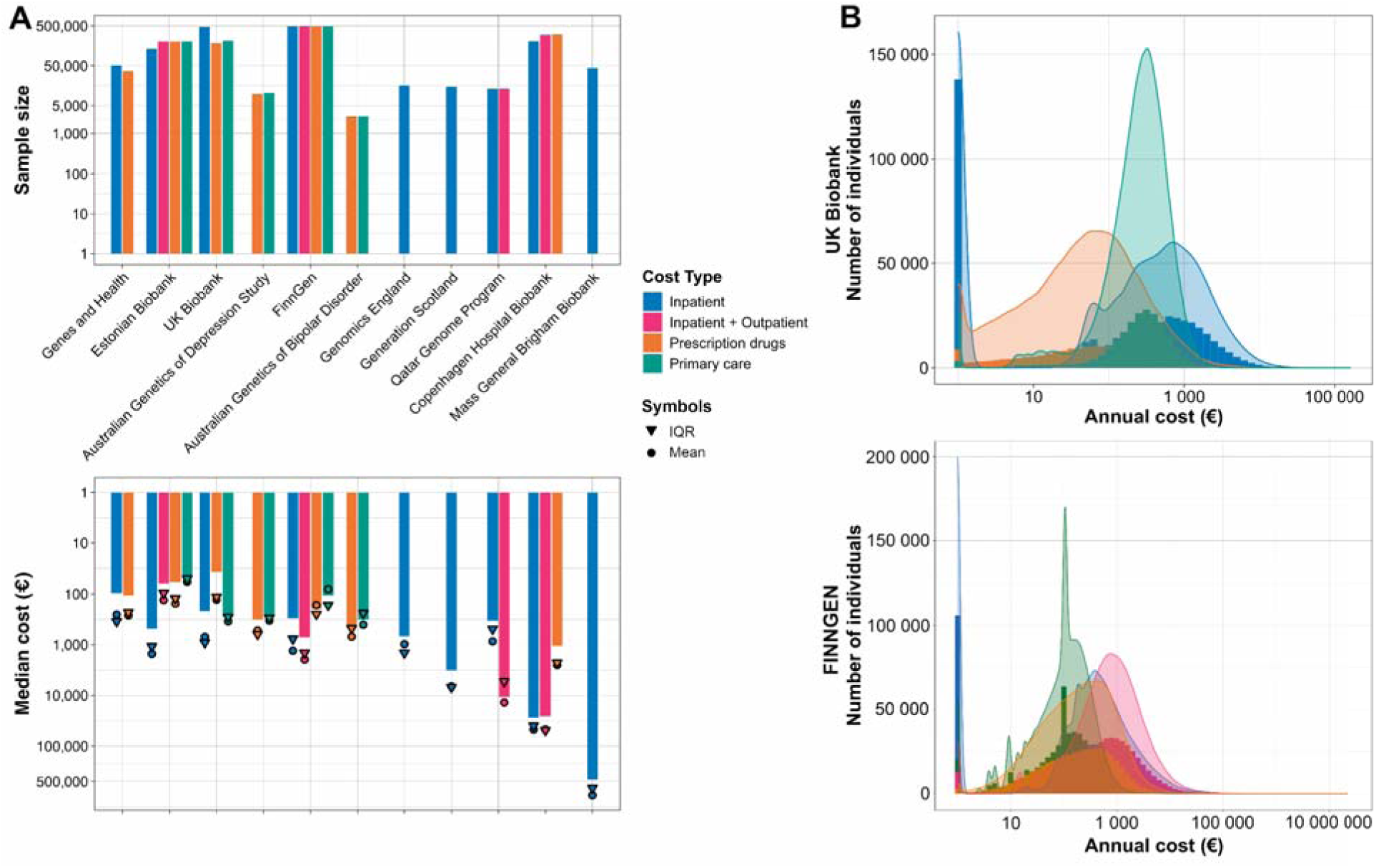
| Cohort characteristics and distribution of healthcare costs. (**A**) Bar plots showing sample sizes (top) and distribution of median healthcare costs (bottom) across 11 contributing cohorts and cost subtypes. Bars are coloured by cost subtype; arrows indicate upper interquantile range (IQR) and circles represent mean values. Both panels are displayed on a logarithmic scale. **(B)** Histograms with density overlays for UK Biobank (top) and FinnGen (bottom), stratified by cost subtype.

We analysed four primary cost phenotypes, derived from electronic health records or administrative databases: (i) inpatient costs (expenditures from overnight hospital stays), N = 1,429,889, (ii) inpatient plus outpatient costs (henceforth stylised as inpatient + outpatient), N = 994,451, (iii) primary care costs (visits to general practitioners and family physicians), N = 914,643, and (iv) prescription costs (costs of medications), N = 1,279,645. These phenotypes are recorded across multiple cohorts, with all four available in two cohorts (Estonian Biobank and FinnGen), and the inpatient cost phenotype being the most commonly recorded **(Figure 1A)**. All cost measures represent expenditures borne by the healthcare system rather than out-of-pocket costs for individuals. Details of phenotype construction and harmonization are provided in the **Methods** and **Supplementary information**.

To facilitate interpretation and comparison across healthcare systems, we analysed the natural logarithm of inflation-adjusted annual costs accrued over each study’s defined follow-up period. Follow-up duration varied across cohorts but was at least 5 years (with the exception of cohorts from Australia) and costs were corrected for inflation to 2019 prices (except for the FinnGen and Mass General Brigham Biobank studies which use 2017 and 2021 prices) (**Supplementary Table 1**, **Supplementary Information**). Individuals with zero costs were assigned a value of 1 prior to log transformation.

Median annual inpatient costs were broadly similar across the three largest European biobanks (UK Biobank, FinnGen, and Estonia Biobank) at ranging from €276 to €482 per person (**Figure 1A**). In contrast, individuals from the Copenhagen Hospital Biobank (CHB, Denmark) and Mass General Brigham Biobank (MGBB, United States) exhibited substantially higher median costs. This difference reflects cohort-specific recruitment strategies and systemic cost variation: CHB participants were recruited during inpatient hospital stays, capturing a less healthy population, while higher costs in MGBB stem from both recruitment bias of hospital patients but also higher price levels in the U.S. healthcare system.

Across all studies and cost phenotypes, cost distributions showed consistent patterns, as exemplified by the two largest studies: FinnGen and UK Biobank (**Figure 1B**). Costs followed a Tweedie distribution characterized by a peak near zero, corresponding to healthy individuals with minimal or zero healthcare use, and a long right-skewed tail representing a small proportion of individuals incurring disproportionately high expenditures. This feature was observed across the majority of cohorts (**Supplementary Figure 1A-1I**) for every cost category (exceptions include hospital-based cohorts such as CHB, which had no zero-cost individuals). Prescription and primary care costs generally exhibited far fewer zero values than inpatient costs, consistent with most individuals receiving at least one prescription or visiting a GP in the absence of hospitalization. Some expected demographic trends were also observed: all cost types increased with age and men had slightly higher inpatient costs than women while women had slightly higher prescription costs than men (**Supplementary Table 2**).

### Genetic signals associated with healthcare costs

To understand the genetic architecture of healthcare costs, we conducted GWAS meta-analyses followed by conditional analyses using GCTA-COJO^20^. We identified the highest number of conditionally independent SNPs (P-value < 5×10^−8^) for prescription drug costs (198), followed by inpatient + outpatient costs (72), primary care costs (57) and inpatient costs (53) (**Figure 2A, Supplementary Figure 2, Supplementary Table 3**). Additionally, GWAS meta-analyses were conducted for each phenotype stratified into age and sex groups (**Supplementary Table 4)**. By applying positional windows of 500 kb around each significant SNP, we grouped the 380 conditionally independent signals into 248 distinct loci across the four healthcare cost phenotypes. Each locus was then assigned to its nearest gene (**Supplementary Table 5)**. Among genome-wide significant variants, effect sizes are generally modest. For annual inpatient costs, they range from a 1.99% decrease to a 2.64% increase per additional effect allele, with one variant (intronic to *TTC28,* and in LD with a frameshift mutation in *CHEK2*) showing a 6.94% increase (**Supplementary Table 3, 4**).

**Figure 2.**
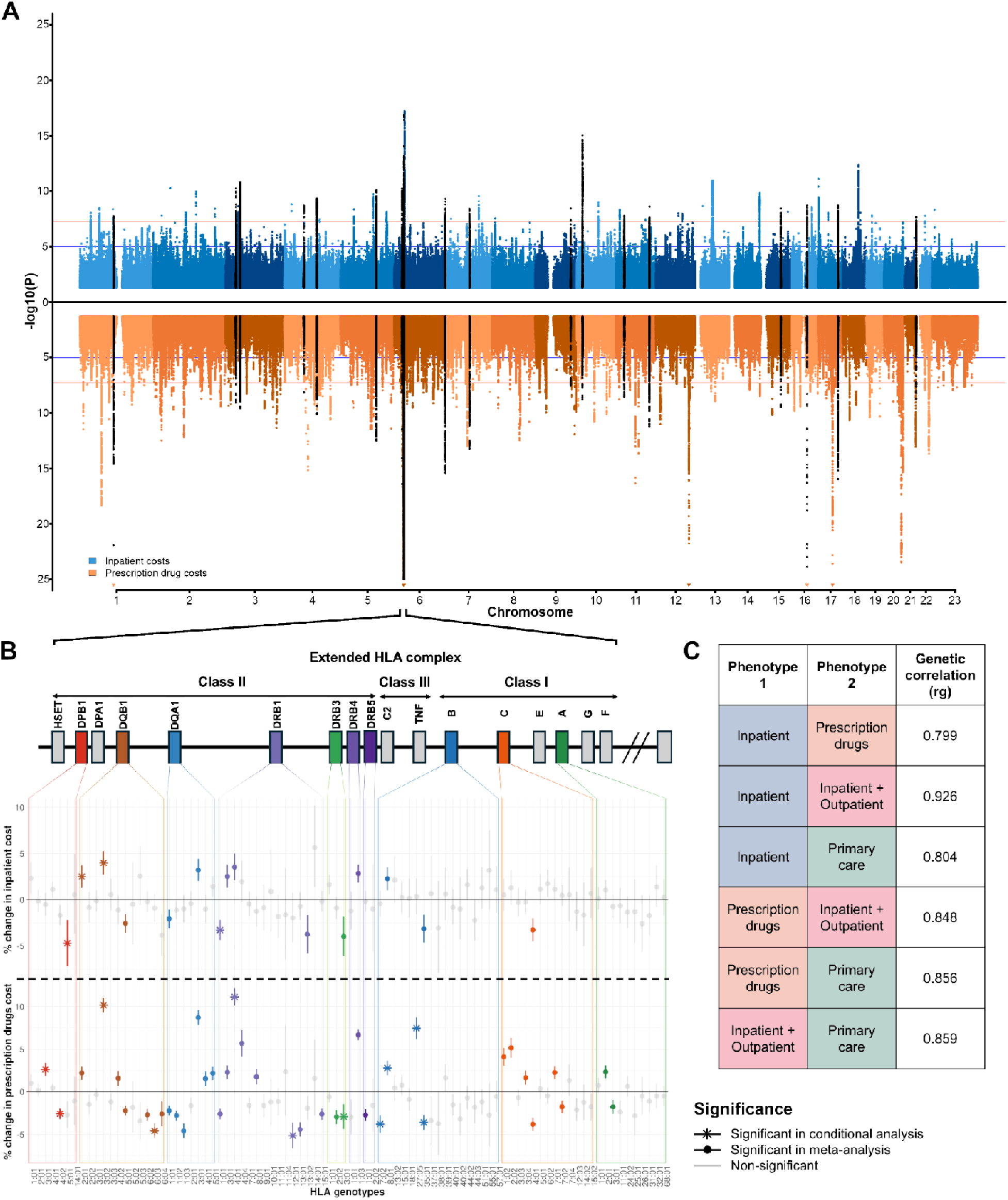
| Genome-wide association of healthcare costs and HLA fine-mapping. (**A**) Miami plot of GWAS results for inpatient (blue, *N* = 1,429,889) and prescription drug costs (orange, *N* = 1,279,645). Loci shared between both phenotypes are shown in black. **(B)** Schematic of the HLA region on chromosome 6 with allele-level association results from meta-analysis of UK Biobank and FinnGen. Effect sizes are expressed as percentage change in costs per allele, shown separately for inpatient (top) and prescription costs (bottom). Coloured points indicate Bonferroni-corrected (*P < 0.05/88*) significant associations, and stars denote variants remaining significant after stepwise conditional analysis. **(C)** Table of genetic correlations between the four cost phenotypes. All correlation pairs shown were significantly different from both zero and one.

Strong positive genetic correlations were observed between all four cost phenotypes (*rg* = 0.80–0.93; **Figure 2C, Supplementary Table 6**). These correlations were significantly different from both zero and one, indicating that while the phenotypes share substantial genetic architecture, they are not identical. Consistent with this, 53 of the 248 loci were genome-wide significantly associated with more than one phenotype and always with the same direction of effect (**Supplementary Figure 3**).

The locus with the most significant signals in all four phenotypes was the HLA locus, consistent with its complex and pleiotropic disease associations^21^. To further characterize its contribution, we tested associations between 88 imputed HLA common alleles and both inpatient and prescription drug costs in the UK Biobank and FinnGen, followed by the meta-analysis of both cohorts (**Supplementary Table 7**). We identified 15 HLA alleles significantly associated with inpatient costs and 39 with prescription costs (Bonferroni-corrected P-value < 0.05/88). Stepwise conditional analyses retained 4 and 11 independent alleles for the two phenotypes, respectively (**Figure 2B**). Notably, the DQB1*03:02 and DRB1*04:01 HLA alleles showed the strongest positive effects in both phenotypes, being independently and respectively associated with 3.98% [CI: 2.7 – 5.2] higher annual inpatient and 11.07% [CI: 10 – 12.2] higher annual prescription costs. DQB1*03:02 confers increased risk for celiac disease and type 1 diabetes, contributing to a significant proportion of all genetic risk for these diseases^22–26^, while DRB1*04:01 also increases risk for type 1 diabetes along with rheumatoid arthritis^27–30^. Conversely, DPB1*05:01 was linked to a 4.9% [CI: –7.7 – –2.2] reduction in inpatient costs which has previously been shown to be associated with spontaneous clearance of the hepatitis B virus in Caucasian populations^31^, however this is unlikely to fully explain the observed reduction in inpatient costs.

### Heterogeneity across studies and by age and sex

We estimated genetic correlations between cohorts to assess the consistency of genetic signals across healthcare systems and cost phenotype definitions (**Supplementary Tables 8-11**). Genetic correlations could not be estimated for all study pairs due to the low heritability of certain cost phenotypes in specific cohorts. For inpatient costs, the phenotype available in most studies, genetic correlations exceeded 50% in 52 of 55 study pairs (**Supplementary Table 8**), indicating substantial concordance of genetic signals across studies.

Consistent with this, we assessed heterogeneity in effect sizes across studies for the 380 conditionally independent genome-wide significant lead variants using Cochran’s Q statistic. Significant heterogeneity (P < 0.05; FDR correction; **Supplementary Table 12**) was observed for 24 variants, which were primarily located in the HLA region. Outside this region, there was little evidence of meaningful heterogeneity in effect sizes across studies.

We next examined heterogeneity across age groups and sex by performing additional GWAS stratified by sex and by age groups (5–18, 19–35, 36–55, 56–75, and ≥76 years). We excluded 62 of the 380 variants, due to their location in the HLA region and focused on 318 non-HLA independent signals identified in the four primary cost-phenotype analyses. Sex differences were assessed using Cochran’s Q test, while age-related heterogeneity was evaluated as using linear meta-regression across age groups. We observed several general patterns (**Figure 3**; **Supplementary Table 12**). Effect sizes for loci associated with prescription drug costs were generally larger in males than in females, whereas the opposite trend was observed for inpatient costs; genetic effects, particularly for prescription costs, also tended to increase with age, partly reflecting our greater statistical power to detect genome-wide significant associations in older individuals. Among the 318 variants, 39 showed nominal evidence of sex heterogeneity (P < 0.05) and 53 of age-related heterogeneity (P < 0.05). After controlling for multiple testing using a 5% false discovery rate, 2 sex-heterogeneous and 10 age-heterogeneous variants remained significant.

**Figure 3.**
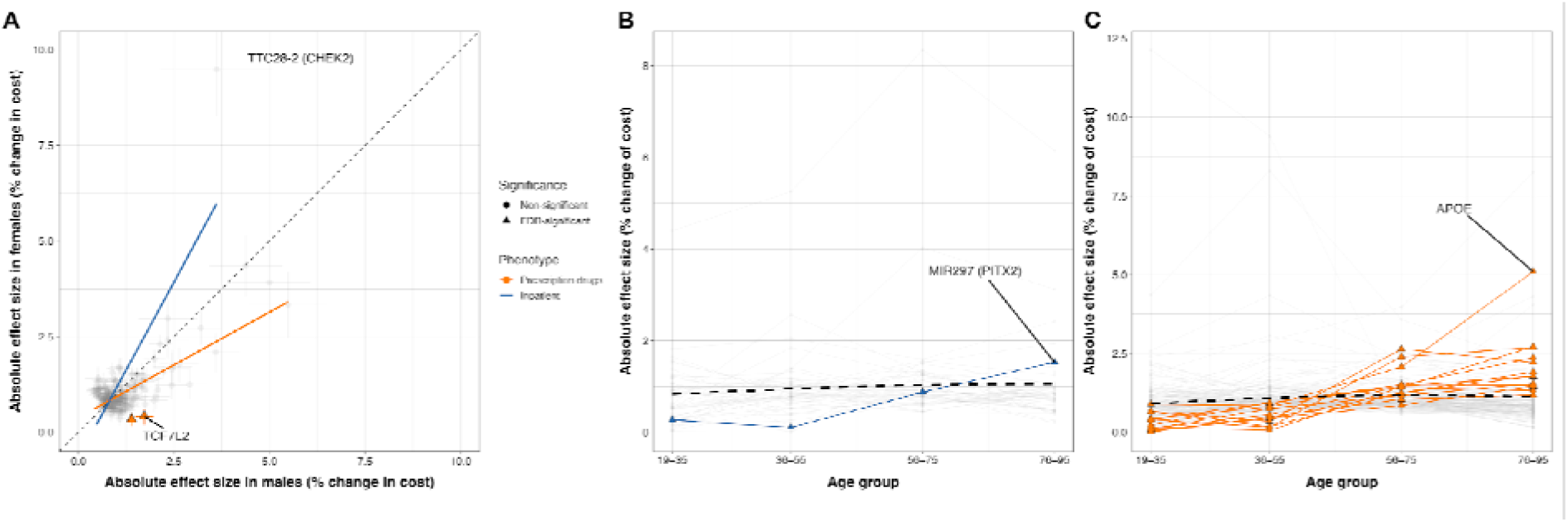
| Heterogeneity of genetic associations by age and sex. (**A**) Absolute effect sizes of 318 non-HLA sentinel SNPs in males versus females for inpatient and prescription cost phenotypes. Points show the absolute percentage change in annual costs per effect allele; greyed-out circles indicate SNPs with non-FDR significant sex heterogeneity by Cochran’s Q-test, and coloured triangles indicate SNPs that pass FDR correction for sex heterogeneity. **(B–C)** Absolute effect sizes of the same SNPs across age bins for inpatient **(B)** and prescription **(C)** costs. Points again denote the absolute percentage change in costs within each age group, with circles marking SNPs showing non-FDR significant trends and triangles marking SNPs that pass FDR correction for age-related heterogeneity.

Rs55972445, an intronic variant in *TCF7L2*, showed FDR-significant sex heterogeneity with a stronger effect in men (1.7% [1.5–1.9] increase per allele) than in women (0.4% [0.2–0.6]). Given the well-established association between *TCF7L2* and type 2 diabetes^32^, this difference may reflect the higher incidence of type 2 diabetes in men. Rs62237617 at the *TTC28* (*CHEK2*) locus showed nominally significant sex heterogeneity for inpatient costs (Q-test P-value = 0.004). The variant, which is in perfect linkage disequilibrium with a *CHEK2* frameshift variant (rs555607708), was associated with a larger increase in inpatient costs in women (9.5% [8.2-10.8] increase per allele) than in men (3.6% [2.0-5.1]), consistent with the elevated breast cancer risk conferred by *CHEK2* variants in women^33^. The strongest evidence for linear age-related heterogeneity in prescription drug costs was observed for rs1065853 in the APOE locus (P = 2.5 × 10⁻⁸). Effect sizes increased with age, from 0.89% and 0.84% in individuals aged 19–35 and 36–55 years to 2.07% and 5.1% in those aged 56–75 and ≥76 years, respectively, consistent with the established role of APOE in age-related disease phenotypes^34,35^.

### Attributable annual healthcare cost for rare deleterious variants in UK Biobank

We used whole-exome sequencing data from 409,927 UK Biobank participants of European ancestry to estimate annual inpatient healthcare costs associated with rare deleterious coding variants in 89 clinically relevant genes, including genes recommended for the return of incidental findings in clinical sequencing, monogenic diabetes genes, and genes linked to cardiovascular risk factors (**Methods**). We considered the burden (sum of alleles) of rare (MAF < 0.001) deleterious variants within each gene using increasingly inclusive pathogenicity definitions (**Methods, Supplementary Table 13**).

From these analyses, 4 genes, all linked to cancer predisposition, were significantly associated with inpatient costs, when considering putative loss-of-function variant burden (pLOF; defined as carriers of ClinVar/ENIGMA pathogenic variants or LOFTEE high-confidence pLOF variants, P < 0.05; FDR correction): *BRCA1*, *BRCA2*, *MSH2*, and *APC* (**Figure 4A, B, Supplementary Table 13**). A pLOF variant carrier in *BRCA2* had on average 61.2% (95% CI: 37.0-89.7%) higher annual inpatient costs than non-carriers in European ancestry. Concordant with a higher risk of female related cancers, sex stratified analysis revealed that there is a large sex-difference in effect sizes for *BRCA2* mutation (100.0% [59.9%-150.2%] increase in females vs 26.8% [0%-60.6%] increase in males; **(Supplementary Table 14).**

**Figure 4.**
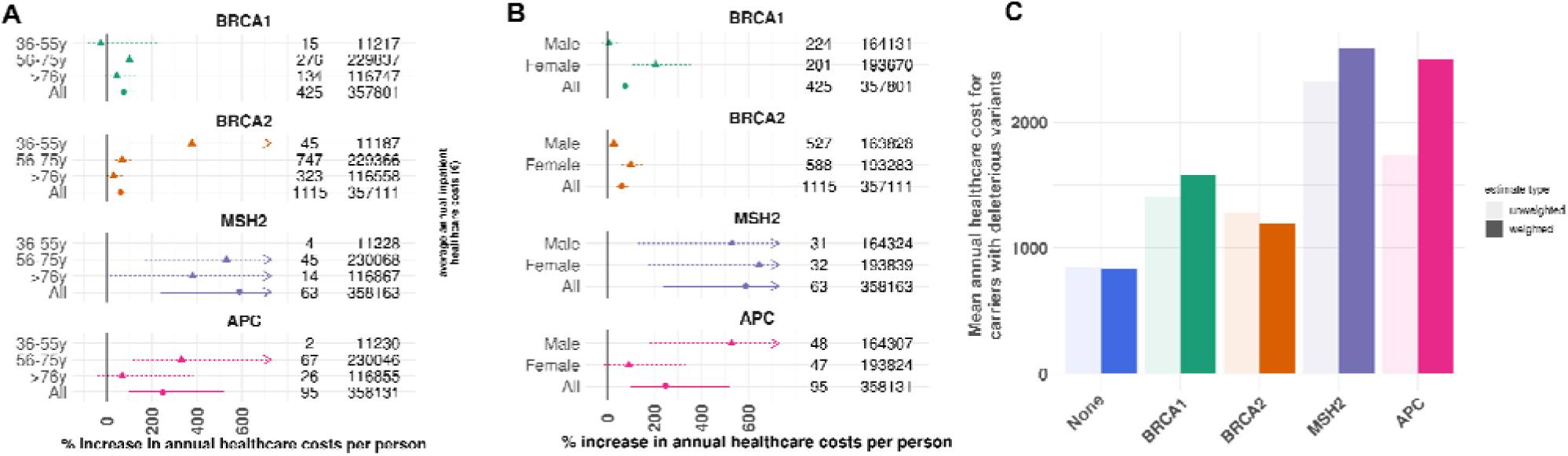
| Rare coding variation in clinically relevant genes and healthcare costs. (**A**) Age-stratified attributable costs. Percent increase in annual inpatient healthcare costs per person for carriers of rare deleterious variants in *BRCA1*, *BRCA2*, *MSH2*, and *APC*, stratified by age group (36–55y, 56–75y, >76y) and overall. Point estimates with 95% confidence intervals are shown; numbers indicate carriers (left column) and non-carriers (right column) in each group. **(B)** Sex-stratified attributable costs. Percent increase in annual inpatient healthcare costs per person for male and female carriers of rare deleterious variants in the same four genes. Numbers indicate carriers and non-carriers in each group. **(C)** Average attributable annual inpatient healthcare costs for carriers of rare deleterious variants, shown overall and separately for *BRCA1*, *BRCA2*, *MSH2*, and *APC*. Bars represent unweighted (light) and probability-weighted (dark) estimates, with error bars denoting standard deviations.

Because UK biobank is a volunteer-based cohort and not fully representative of the UK population, we performed a probability-weighted analysis using external Health Survey for England datasets^36^. After weighting, the estimated mean annual inpatient cost among *BRCA2* pLOF carriers was €1188.5 (SD €1877.2), compared with €831.3 (SD €1635.7) in the general population (**Figure 4C**, **Supplementary Table 15**). Extrapolating this to the national level and assuming the same frequency of *BRCA2* pLOF carriers, the attributable annual inpatient cost in UK adults of European ancestry aged 40–69 was approximately €17.4 million (**Supplementary Table 15**).

To replicate these findings, we performed analogous analyses in three additional cohorts including individuals of diverse ancestries: UK Biobank participants of Central and South Asian ancestry (N = 9,790), Genomics England (European ancestry) (N = 13,618), and East London Genes & Health (Central and South Asian ancestry) (N = 25,904). An inverse-variance weighted meta-analysis across cohorts, showing consistent direction of effect for *BRCA1*, *BRCA2*, *MSH2*, and *APC* (**Supplementary Table 16**), although associations did not reach statistical significance, likely due to the small number of variant carriers.

### Identification of phenotypic drivers of genetic associations with healthcare costs

To elucidate the diseases and traits underlying the association between genetic variation and costs we performed an explorative phenotype-wide association study to find previously discovered GWAS associations of our reported sentinel SNPs. Of the 380 conditionally independent sentinel SNPs, 349 had evidence of previous associations with multiple different traits based on the FinnGen annotation browser. This resource collates GWAS associations from GWAS catalogue, OpenTarget Genetics (version 22.09) and publicly GWAS summary statistics combining UK Biobank, FinnGen (Release 13) and the Million Veterans Programme (**Supplementary Table 17**). Of the remaining 31 SNPs, 17 were in high LD (R2 > 0.8) with a SNP which also had evidence of a previously discovered GWAS association (**Supplementary Table 18**).

This left 14 conditionally independent lead SNPs with no previously reported phenotype associations in our initial annotation resources. After manual curation using a recent release of the Open Targets Genetics platform (version 25.12), 5 of the 14 SNPs mapped to a credible set for at least one other disease or trait (**Supplementary Table 19**). The remaining 9 SNPs had no previously reported genome-wide significant associations. Nevertheless, 7 of these 9 SNPs showed nominal evidence of association with disease endpoints in FinnGen R13 (Phewas significance threshold: P < 0.00016). For example, rs72988527 showed nominal association with autoimmune disease endpoints (excluding thyroid-related diagnoses) and was associated with higher inpatient and outpatient costs. rs4147268 in *ESRRG* was associated with higher prescription drug costs and showed nominal association with depression, and rs248618 showed nominal associations with atrial fibrillation. In contrast, rs56115704 and rs10234261 showed no detectable associations with any disease endpoints, even at nominal thresholds, in FinnGen. This pattern suggests that these loci may influence costs through mechanisms not well captured by current disease phenotypes, or that relevant clinical endpoints remain underpowered in the available resources.

To test whether cost loci share causal variants with established biomedical traits, we conducted targeted colocalization analyses between loci associated with the four cost phenotypes and GWAS results from a curated set of 45 traits spanning common diseases, risk factors, and laboratory measures (**Supplementary Table 20**). Using COLOC under a single-causal-variant assumption, we identified 1105 colocalized signals (PP4 ≥ 0.8) across 295 distinct variants in 194 loci (**Supplementary Table 21**). Loci associated with prescription drug costs exhibited the highest number of colocalizing signals (n = 579), consistent with this phenotype also yielding the largest number of genome-wide loci, whereas each of the remaining cost phenotypes contributed substantially fewer (n = 163-191). The traits with the greatest number of colocalizing loci included hypertension, asthma, coronary atherosclerosis, joint pain and major depression (**Figure 5A**). These highly prevalent, often chronic and comorbid conditions drive sustained medical management, leading to cumulative healthcare costs associated with long-term monitoring and medication use. However, these traits are also among the best-powered GWASs, which likely contributes to increased power for detecting colocalization.

**Figure 5.**
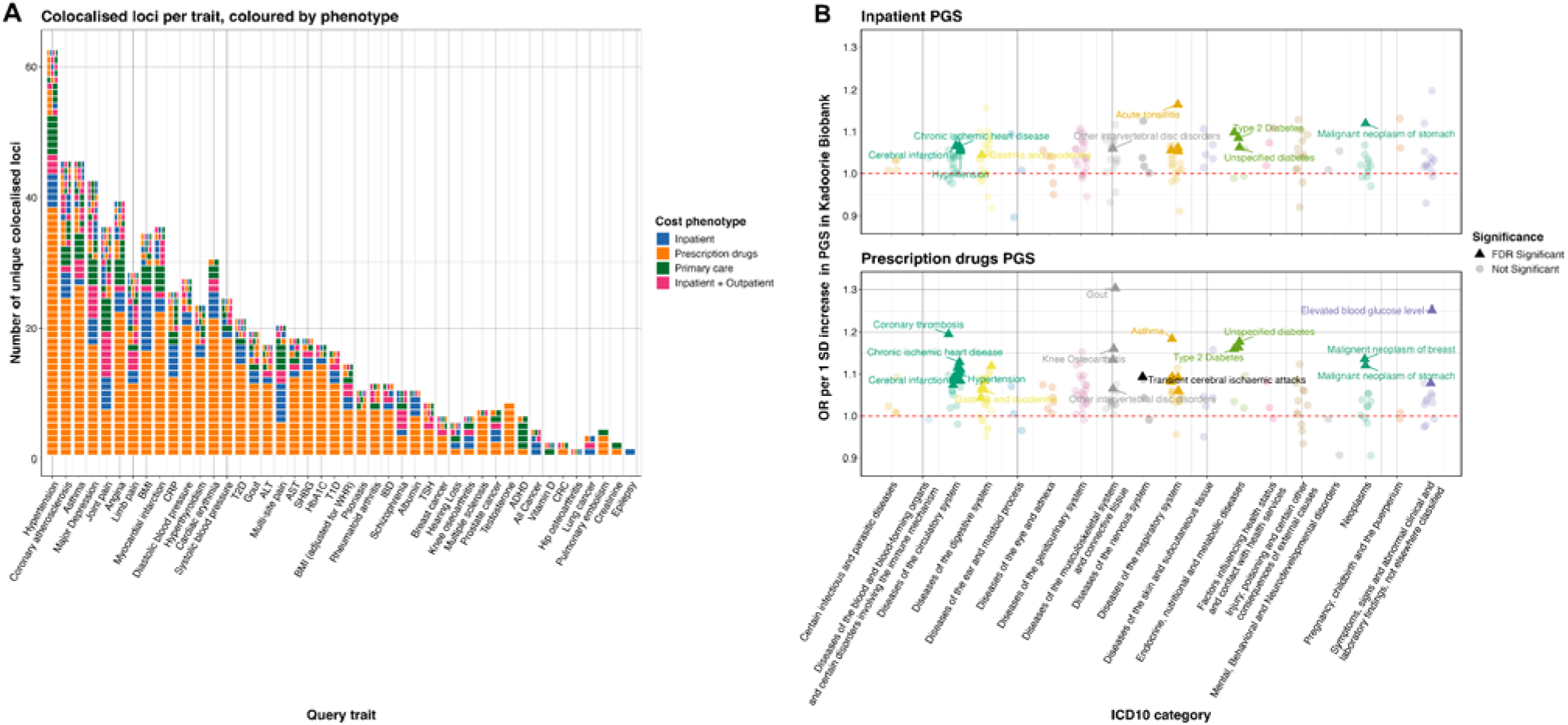
| Colocalisation of cost-associated loci with complex traits and disease outcomes. (**A**) Tile plot showing colocalisation between independent autosomal loci identified in the four healthcare cost GWAS and 45 selected complex traits. The x-axis lists the queried traits, ordered by the total number of colocalising loci, and the y-axis indicates the number of unique colocalised loci. When the same locus colocalises with a given trait across multiple cost phenotypes, the corresponding tile is partitioned by colour according to the cost phenotype. **(B)** Associations between PGS for inpatient costs (top) and prescription drug costs (bottom) and first occurrence of 172 different disease events defined by ICD-10 codes in the China Kadoorie Biobank. Effect estimates are shown per one standard deviation increase in PGS. Associations surviving FDR correction are highlighted.

The variant showing the strongest evidence of colocalization across phenotypes was rs3184504, a missense variant in *ATXN2*/*SH2B3*. This variant has been robustly associated with hypertension, autoimmune disorders, and a broad range of immune and hematopoietic traits^37–39^. *SH2B3* encodes an adaptor protein that modulates multiple signaling pathways and plays a central role in hematopoiesis, inflammation, and cell migration, and has been shown to act as a negative regulator of cell proliferation^40^.

Five loci had colocalisations which were shared between all four cost phenotypes: *METTL15*, *MAD1L1*, *FTO*, *BPTF*, and *ARPP21*. Notably, variants in the *METTL15* locus colocalized with limb and joint pain across all cost categories, consistent with increased expenditures spanning prescriptions, inpatient care, primary care, and combined inpatient– outpatient settings. *FTO* exhibited broad pleiotropy with colocalisations in BMI, breast cancer, osteoarthritis and several biomarkers, potentially reflecting its established effect on body mass index and downstream cardiometabolic risk^41^.

To investigate the disease pathways mediating the relationship between genetic liability and healthcare utilization, we constructed polygenic scores (PGS) for each cost phenotype and tested their associations with the first occurrence of 172 diseases in 82,389 participants from the China Kadoorie Biobank (**Figure 5B**). As an external cohort, this provides independent validation with longitudinal disease ascertainment from electronic health records. The inpatient cost PGS was significantly associated, after FDR-correction, with multiple cardiometabolic diseases, specifically ischemic heart disease, stroke and hypertension, as well as malignant stomach neoplasms (**Supplementary Table 22**). The drug cost PGS showed stronger and more diverse disease associations, with significantly larger effects observed for diabetes, infectious diseases, and asthma. The largest association was observed for gout (OR = 1.3 [1.25-1.35]), consistent with the broad colocalization signal between gout and drug prescription costs (with 17 loci colocalising between prescriptions and gout), suggesting that the association between gout and prescription costs is supported by multiple independent genetic signals rather than a single or a few pleiotropic loci.

### Population and within-family polygenic prediction of healthcare costs

We evaluated the predictive performance of the PGS in an external cohort not included in the GWAS meta-analysis, the Netherlands Twin Registry (NTR, N = 15,440), which includes high-quality individual-level healthcare cost data obtained directly from Statistics Netherlands. We observed significant linear associations between the prescription drug PGS and drug expenditures, and between the inpatient cost PGS and inpatient + outpatient expenditures (**Figure 6A, B**).

**Figure 6.**
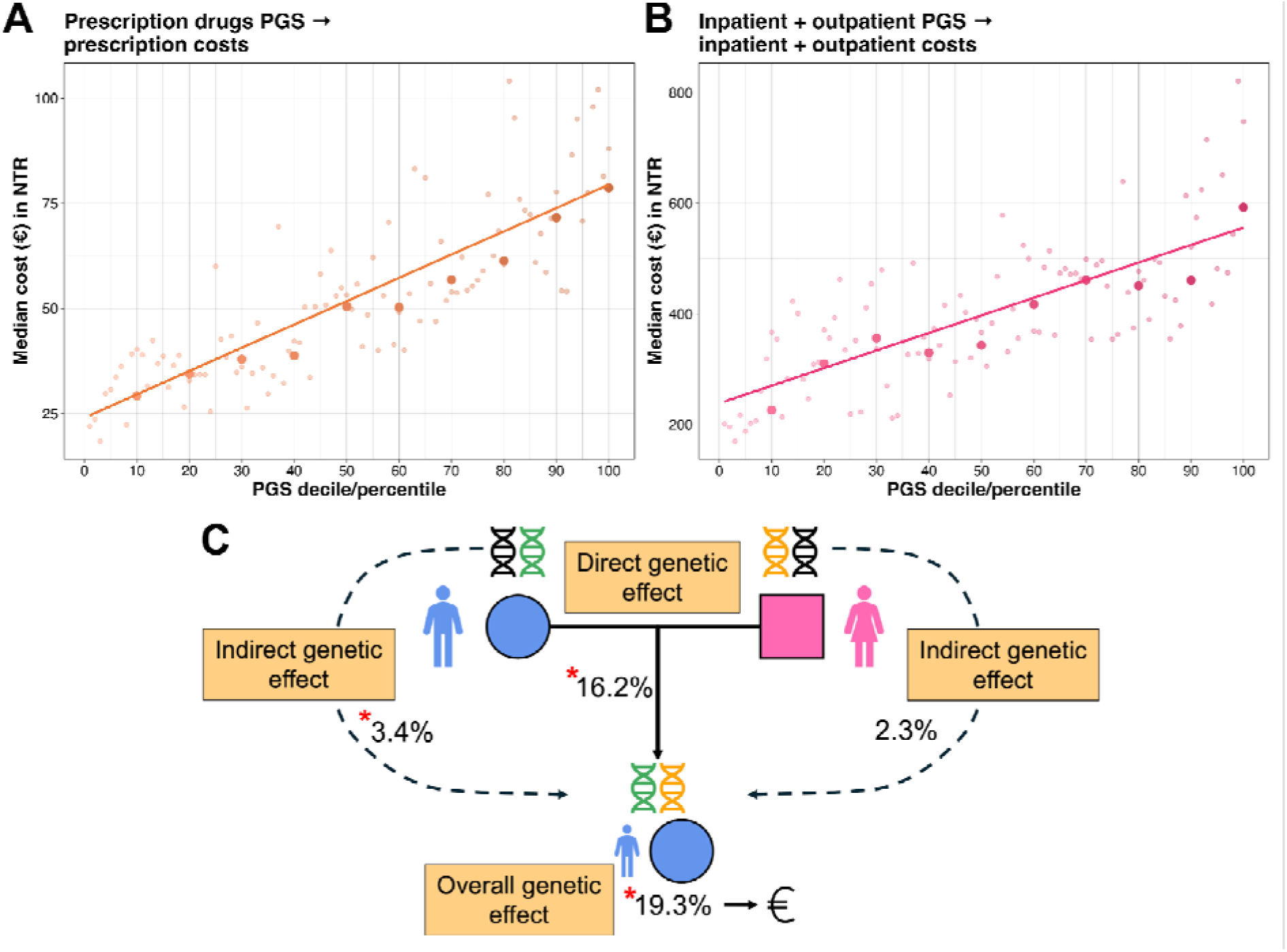
| Polygenic prediction of healthcare costs. Associations between the prescription drug PGS and drug expenditures (**A**), and between the inpatient cost PGS and inpatient + outpatient expenditures (**B**) in the Netherlands Twin Register (NTR, N=15,440). Smaller, transparent dots represent costs by PGS centiles, while larger solid dots represents PGS deciles. (**C**) PGS transmission analysis in 181,686 FinnGen trios, comparing effects of offspring PGS, maternal PGS, and paternal PGS on inpatient costs.

Individuals, within NTR, in the top decile of the prescriptions PGS have a median annual cost of €78.64 compared with €29.24 among those in the lowest decile, corresponding to an 168% higher annual cost. For inpatient + outpatient costs, the corresponding medians were €592.72 versus €225.90 (162% higher). The PGS for prescriptions demonstrated the highest predictive power, explaining 1.9% of the variance (R²) in prescriptions (**Supplementary Table 23**).

Despite successful external replication, PGS for complex phenotypes can still capture population stratification, indirect genetic effects, and other sources of bias^42^. A robust approach to mitigate these biases is to test PGS-phenotype associations within families^43^. We therefore recalculated the PGS for inpatient costs after excluding FinnGen from the meta-analysis and tested its association in 181,686 parent–offspring trios from FinnGen (**Supplementary Table 24**), where genotypes were either directly measured or imputed using SNIPAR^44^ (**Figure 6B**). Without accounting for family structure, one standard deviation increase in the inpatient cost PGS was associated with a 19.3% higher annual inpatient cost (P < 1 x 10^−5^, Wald test). After adjusting for paternal and maternal PGS, this effect attenuated somewhat to 16.2% but remained strongly significant (P < 1 x 10^−5^, Wald test). We also detected a significant association between paternal PGS and annual inpatient costs contributed by the offspring (3.4% increase, P = 0.008), suggesting that non-transmitted paternal alleles, possibly reflecting indirect genetic effects, contribute to variation in offspring healthcare expenditure.

Together, these findings indicate that polygenic influences on healthcare costs are replicable in an external healthcare system and largely reflect direct genetic effects rather than confounding from population structure or shared family environment.

### Established polygenic scores have a substantial impact on healthcare costs in FinnGen

We evaluated the association between 966 PGS (not derived using FinnGen data) obtained from the PGS Catalog and healthcare costs in the FinnGen study^45,46^. These PGS capture genetic liability across a broad spectrum of traits and diseases. We identified 1,512 significant PGS-phenotype associations, with 489 PGS significantly associated with at least one healthcare cost phenotype (P < 0.05/966), indicating a pervasive contribution of established polygenic risk to variation in healthcare expenditures **(Supplementary Table 25**). For example, higher genetic liability for abdominal pain (PGS002096) was associated with a 14% [13.6-14.5%] increase in inpatient healthcare costs per one standard deviation (SD) increase in the PGS (**Figure 7A**). Conversely, the PGS for college education, (PGS002319) was strongly associated with lower healthcare costs across all four cost phenotypes (−5.1% to −13% per standard deviation increase in PGS). Together with associations observed for PGSs indexing behavioural and life-course traits, such as age at smoking initiation and age at first live birth, these findings likely reflect the cumulative impact of socioeconomic position and health-related behaviours on healthcare utilisation.

**Figure 7.**
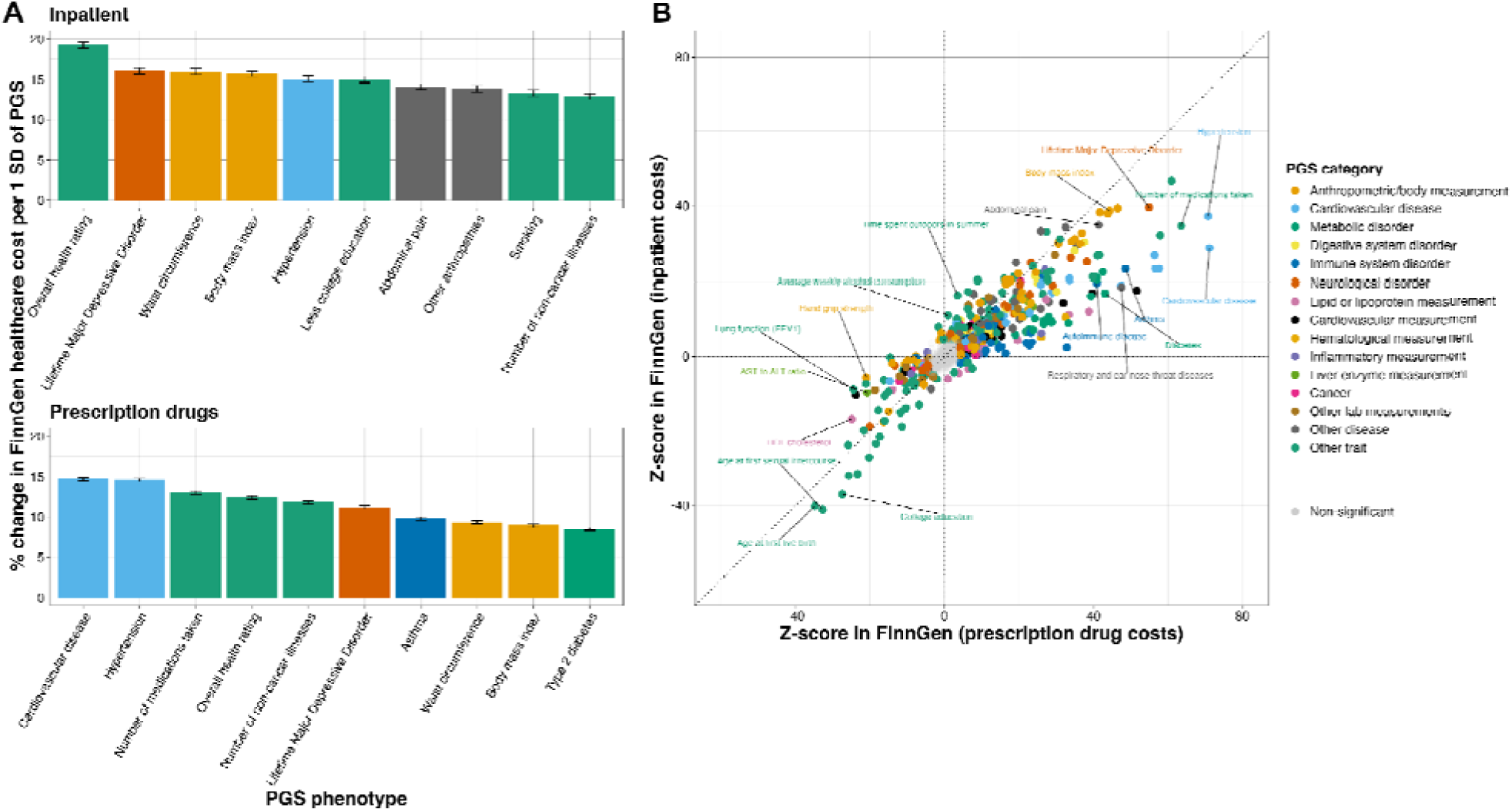
| Associations between established scores and inpatient and prescription drug costs. (**A**) Selected PGS showing the strongest associations with inpatient costs (top) and prescription drug costs (bottom). Bars represent the percentage change in healthcare costs per one SD increase in the corresponding PGS, with error bars indicating 95% confidence intervals. **(B)** Comparison of PGS associations with inpatient and prescription drug costs. Each point represents a single PGS, plotted by its Z-score for the association with inpatient costs (x-axis) and Z-score for the association with prescription drug costs (y-axis). Points are coloured by PGS category. PGS showing a statistically significant difference in effect size between inpatient and prescription drug costs (P < 0.05/966) are highlighted.

Overall, PGS effects were more pronounced for prescription costs than for inpatient drug costs, although notable exceptions were observed (**Figure 7B**). For example, the PGS for overall health rating (PGS002000) showed a larger association with inpatient costs (19.2% [18.8-19.7%] per SD) than with prescription costs (12.5% [12.2-12.7%]) potentially reflecting hospitalised individuals generally having worse health than those obtaining prescriptions (**Figure 7B**).

To assess the joint contribution of the 966 genetic risk across traits, we included all PGS simultaneously in a single multivariable model. Collectively, all the PGS explained a 1.4% and 2.5% of the variance (R2) (**Supplementary Table 26**) in inpatient and prescription drug costs, respectively, beyond a baseline model including age, sex, and ten genetic principal components. As a comparison, the PGS for costs, after excluding FinnGen from the meta-analysis, explained, in FinnGen, 0.5% and 1.4% of the variance in inpatient and drug costs, respectively (**Supplementary Table 26**)

Together, these findings show that established polygenic risk for diseases and their risk factors captures a measurable component of healthcare cost variation and can inform cost prediction to an extent which even exceeds that achieved by PGS derived directly for healthcare cost phenotypes.

## Discussion

Healthcare utilization varies markedly between individuals, resulting in substantial heterogeneity in healthcare costs. Here, we show that a measurable and systematic component of this variation is attributable to genetic differences between individuals. By meta-analysing data from 11 studies across different healthcare systems, we identified hundreds of genetic variants robustly associated with healthcare costs, providing a large-scale map of the genetic architecture underlying healthcare expenditure.

At the level of common variation, individual variants typically had modest effects, increasing annual inpatient costs by approximately ∼1-2%. This pattern mirrors the small effect of common variants on complex diseases which lead to healthcare utilization. SNPs in the MHC/HLA-region showed the strongest and most significant associations with healthcare costs, with multiple independent HLA alleles associated with ∼5–10% higher inpatient and drug prescription costs in both FinnGen and UK Biobank. Given the extensive pleiotropy of HLA variation across immune and inflammatory diseases, and the relatively high frequency of these alleles in individuals of European ancestry (e.g. ∼10.5% of individuals in UK Biobank have at least one copy of the HLA DRB1*4:01 allele), this region emerges as a major contributor to population-level variation in healthcare costs. At the same time, effect sizes in the HLA region showed substantial heterogeneity across studies, reflecting well-known differences in HLA allele frequencies across populations, and highlighting challenges for generalization. Outside the HLA region, genetic correlation and heterogeneity analyses indicated broadly consistent effects across cohorts.

In contrast to the modest effects of common variants, rare deleterious mutations in clinically actionable genes had pronounced individual-level consequences. Carriers of loss-of-function variants in *BRCA1*, *BRCA2*, *MSH2*, and *APC* experienced more than a two-fold increase in annual inpatient costs, reflecting the substantial healthcare burden associated with cancer predisposition. Extrapolating and re-weighting these effects to reflect the entire UK population aged between 40 to 69 years old (and of European ancestry), we estimate that *BRCA2* loss-of-function carriers alone account for an additional €17.4 million (£15.1 million) in annual inpatient costs. Importantly, these estimates capture cumulative healthcare expenditures incurred across conditions and over time, rather than costs attributable to a single cancer diagnosis. As such, they provide empirically grounded inputs for decision-analytic models, including those evaluating screening and preventive interventions, which often rely on simplified assumptions about disease incidence or treatment costs.

Beyond cancer and autoimmune disease, a large fraction of the genetic signal underlying healthcare costs mapped to cardiometabolic risk factors, adiposity, pain sensitivity, and inflammatory processes. These patterns were reinforced by analyses in the China Kadoorie Biobank, where polygenic scores for inpatient healthcare costs showed their strongest associations with cardiometabolic disease outcomes, despite substantial differences in ancestry and healthcare systems. The prominent contribution of pain-related genetic variation is consistent with prior reports showing that genetic factors influencing pain are major drivers of loss in disability-adjusted life years^47^. Age– and sex-stratified analyses further revealed loci with demographic-specific effects for both common and rare variation, reflecting differences in disease prevalence, clinical presentation, and healthcare use across the life course and between sexes.

Despite this heterogeneity, polygenic scores derived from our meta-analysis showed robust associations with high-quality healthcare cost data in an independent cohort from the Netherlands, representing a healthcare system not included in the discovery analyses. These associations persisted in within-family analyses, indicating that population stratification and indirect genetic effects are unlikely to account for the observed findings. Notably, existing polygenic scores for diseases and health-related traits explained a larger proportion of variance in healthcare costs as polygenic scores derived directly for healthcare cost phenotypes.

This study has several limitations. First, definitions of healthcare costs, study populations, and reimbursement structures varied substantially across cohorts. As a result, large biobanks such as FinnGen and UK Biobank disproportionately influenced the meta-analysis, limiting generalizability to other settings. Costs were log-transformed, with an offset added for zero-cost observations. While this approach is unlikely to affect locus discovery, it complicates interpretation of absolute effect sizes on the raw monetary scale. Moreover, most participants were of European ancestry, and findings may not fully extend to other populations or healthcare systems. Although we observed significant genetic correlations between European cohorts and the U.S.-based Mass General Brigham Biobank, generalization to the U.S. context remains uncertain, particularly as inpatient costs in that cohort were approximated using Medicare-like reimbursement rates.

Second, extrapolation of rare-variant effects to the general UK population assumes that the frequency of deleterious variants in UK Biobank reflects that of the broader population, an assumption likely violated, especially for non-European ancestries.

Third, our results reflect healthcare practices and disease patterns during the study period, which included the COVID-19 pandemic in several cohorts. Consequently, these findings may not generalize to future healthcare costs, which may be influenced by changes in clinical management, screening practices, or demographic trends. We include a companion **FAQ document** to help different audiences interpret our findings.

In conclusion, our results demonstrate that healthcare costs have a measurable genetic architecture shaped by both widespread pleiotropic common variation and clinically-relevant rare high-impact mutations. By linking genetic risk to healthcare expenditure this work provides a foundation for integrating human genetics into health economics, preventive strategies, and population-level screening.

## Methods

### Contributing studies

Description of each contributing study is provided in the **Supplementary Information**. All individuals were recruited following local ethics, legislation requirements and informed consent. Information about ethics approval and genotyping for each cohort is found in **Supplementary Table 1**. Meta-data for the age, sex and cost distributions for each contributing study is found in **Supplementary Table 2**. In total 11 different studies contributed GWAS summary statistics for meta-analysis in at least one of the four primary cost phenotypes. For inclusion, individuals had to have a minimum of 5 years of follow-up (with the exception of the Australian studies which had 4.5 years of cost data due to data-retention laws), and any study had to have a minimum of 2,000 individuals (and at least 200 individuals for each stratified analysis).

### Phenotype definitions

Four primary cost phenotypes were examined, with each cost being the cost to the healthcare system and not the individual: inpatient costs (costs derived from an overnight stay in hospital), prescription costs (costs of various prescriptions to individuals), primary care costs (costs associated with visits to general practitioners outside of hospital), inpatient + outpatient costs (included as an additional phenotype as most cohorts could not provide outpatient costing data). Final costs for each individual across a given period were divided by the number of days of follow-up, multiplied by 365.25 to give yearly costs. For all cohorts except FinnGen and MGBB, costs were converted to 2019 prices using consumer price indices (e.g. for UK Biobank the Bank of England inflation calculator^48^). Costs for FinnGen and MGBB were instead left in 2017 and 2021 prices, respectively. The natural log of the yearly costs were then used for the final phenotype (**Equation 1**).

Costs were derived by each cohort using multiple different methodologies, fully explained in the **Supplementary Information**.

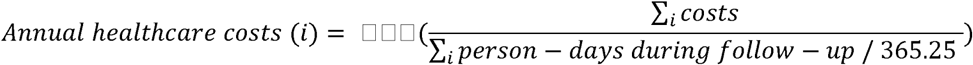

**Equation 1 | Yearly healthcare cost equation**

### Genome-wide association studies (GWAS)

Each cohort performed SNP-level quality control, retaining variants with a call rate ≥ 95% and in Hardy–Weinberg equilibrium, prior to conducting genome-wide association analyses using mixed-models to account for relatedness within cohorts. No MAF or INFO score filter was applied at the GWAS stage, and instead was applied after conducting meta-analysis. Across all 11 cohorts a total of 124,408,332 variants were analysed (the large number primarily driven by results from QGP and GE which use whole-genome sequenced, WGS, variants for their analysis).

Each cohort performed genotyping (or whole genome sequencing), quality control, imputation and analysis independently according to best practices. The methodology used for both UK Biobank and FinnGen, are used as examples of the recommended consortium practices.

We recommended that the GWAS was conducted using REGENIE^49^, or another software capable of carrying out mixed-models which could account for individual relatedness (other such software including BOLT-LMM^50^ and SAIGE^51^). Versions used in each cohort are defined in **Supplementary Table 1**.

It was recommended for cohorts to conduct the analysis using the following covariates: age at the end of follow-up, age at the end of follow-up squared, sex, age multiplied by sex, year of birth and the first 10 or 20 genetic principal components for the cohort (10 for the Australian cohorts, 20 otherwise). Some cohorts also included study-specific covariates such as genotyping batch and array in UK Biobank (**Supplementary Table 1**).

GWAS were also conducted stratified by age and sex. Male and female-specific GWAS were conducted by each cohort, excluding the sex and sex-age interaction covariates for the analysis. For the analyses stratified by age, five age bands were utilised (defined by age at the end of follow-up): 5-18 years old, 19-35 years old, 36-55 years old, 56-75 years old and over 75 years old. A stratified GWAS was only conducted where a minimum of 200 individuals of that strata existed in that cohort.

Summary statistics for each study were examined for potential inflation via manhattan and QQ-plots whilst allele frequencies were compared and alleles aligned with those from gnomAD v3.0. If multiple SNPs were found matching those from gnomAD, the variant with the closest matching frequency to that of gnomAD was selected. Where necessary, e.g. in the case of UK Biobank, SNP positions were lifted over from genome build GRCh37 to build GRCh38, performed using LiftOver.

### Pan-UK Biobank GWAS

To maximise inclusion of individuals across diverse genetic ancestries, GWAS were conducted separately within ancestry-stratified subsets of the UK Biobank. Following the Pan-UK Biobank framework^52^, participants were assigned to one of seven genetic ancestry groups: European (EUR), Central/South Asian (CSA), African (AFR), East Asian (EAS), Middle Eastern (MID), Admixed American (AMR), or unassigned. Each ancestry group was treated as an independent cohort and analysed via GWAS. Cohorts were included as per the study design, in the primary analysis if they comprised at least 2,000 individuals, and in age– and sex-stratified analyses if subgroup sample sizes exceeded 200 individuals. Based on these criteria, individuals of EUR, AFR, CSA, and EAS ancestry were retained for downstream meta-analysis.

### Meta-analysis

Meta-analysis of the GWAS summary statistics was conducted using the METAL analysis software^53^ version 2020-05-05. This was conducted using the following options: SCHEME SAMPLESIZE, for conducting inverse-variance weighted (IVW) meta-analysis methodology based on P-value and direction of effect weighted by sample size GENOMICCONTROL ON, to correct test-statistics for small amounts of population stratification AVERAGEFREQ ON, MINMAXFREQ ON, both to help track any SNPs with differing effect alleles missed in alignment with gnomAD TRACKPOSITIONS ON, to keep hg38 genome positions and ANALYZE HETEROGENEITY, which provides measures of heterogeneity for each SNP in the analysis.

Prior to input into METAL, SNPs with a low imputation INFO score < 0.6 were excluded from the analysis (this did not apply to those studies which provided WGS data). However, no allele frequency filter was applied prior to meta-analysis, with the intention of examining potential rarer variant effects.

### Conditional and joint analysis and sentinel SNP selection

GCTA-COJO^20^ v.1.94.1 was used to conduct conditional and joint analysis to find significant independent loci from each meta-analysis (inpatient, prescription, primary care and inpatient + outpatient costs. For this, the meta-analysis summary statistics were pruned to exclude SNPs with a MAF < 0.001 and for those only present in either one third of the cohorts (or at least two of the cohorts when fewer than 6 cohorts were present) present in each meta-analysis. To provide the LD structure for the analysis, a background panel of 30,000 random, unrelated individuals who identified as white and British from UK Biobank was created.

### Genetic correlation analysis using LDSC

Genetic correlations between cohorts and phenotypes were estimated using linkage disequilibrium score regression (LDSC)^54^. For each phenotype, genome-wide association summary statistics from each cohort were first harmonized and quality-controlled using LDSC’s munge_sumstats procedure, incorporating HapMap3 SNPs and European reference LD scores derived from 1000 Genomes, with squared correlation (r^2^) computed over these SNPs.

Pairwise genetic correlations were then computed using LDSC, both between cohorts for each phenotype and between meta-analysed outputs for each phenotype, using the same European LD reference panel. This procedure allowed estimation of shared genetic architecture across cohorts and between phenotypes, while accounting for differences in allele frequency, LD structure, and SNP coverage.

All correlations were restricted to HapMap3 SNPs with precomputed European LD scores to ensure robust cross-cohort comparisons and minimize bias from differences in SNP density or allele frequency.

Genetic correlations estimated for several cohort-cohort pairs exceeded 1.0. This can occur because the LD Score Regression estimator is not constrained to the interval [−1,1], and the relatively low SNP-heritability of the cost phenotypes results in large standard errors that can inflate estimates above 1.

### HLA meta-analysis and stepwise conditional analysis

Using the three cost phenotypes that overlap between UK Biobank and FinnGen (inpatient, prescription drug and primary care costs) we conducted HLA association analyses in both cohorts. Classical HLA alleles were available at four-digit resolution in each dataset. Analyses were restricted to HLA alleles present in both cohorts and with a minor allele frequency of at least 0.01 in each, resulting in a final set of 88 HLA alleles across 10 HLA genes.

Association testing was performed separately in UK Biobank and FinnGen for each cost phenotype and each HLA allele. In UK Biobank, analyses were restricted to individuals of European genetic ancestry. For each allele-phenotype pair, linear regression models were fitted with the cost phenotype as the outcome and the HLA allele coded additively, adjusting for sex, genotyping batch and array (in UK Biobank specifically), year of birth, age at the end of follow-up, age-squared, age-by-sex interaction, and the first 20 genetic principal components.

Cohort-specific effect estimates and standard errors were combined using IVW meta-analysis. These analyses identified multiple significant HLA associations across all three cost phenotypes.

To identify independent HLA signals within each phenotype, we performed stepwise conditional analyses using a forward-selection framework. For each phenotype, the most significant HLA allele from the meta-analysis was selected as the initial signal. Subsequent regression models were then fitted in both cohorts, conditioning on the previously selected allele(s) and including one additional HLA allele at a time. Conditional effect estimates from UK Biobank and FinnGen were again combined using meta-analysis, and the most significant newly added allele was retained. This process was repeated iteratively, with each new allele added to the existing model, until no further alleles reached statistical significance after multiple testing correction.

At each step of the conditional analysis, statistical significance was assessed using a Bonferroni correction based on the number of HLA alleles tested in that iteration. The procedure terminated when no additional alleles passed the corrected threshold. The final conditional models comprised four independent HLA alleles for inpatient costs, five for primary care costs, and eleven for prescription drug costs, (I.e. the latter requiring eleven sequential rounds of regression and meta-analysis before no further independent signals were detected).

### Rare deleterious variants and gene selection

We evaluated the impact of rare deleterious variants on annual healthcare costs using whole exome sequencing data from the UK Biobank (UKB; n = 409,927 individuals of European ancestry). Genotypes were obtained from UKB provided PLINK files (data field 23158). Variants were annotated using the Ensembl Variant Effect Predictor (VEP; version 109^55^), and high confidence putative loss of function (pLOF) variants were flagged with LOFTEE^56^. Germline variants in BRCA1 and BRCA2 were additionally curated from the ENIGMA consortium^57^ (https://brcaexchange.org/releases, release 58; accessed June 2023). Candidate genes were defined a priori from three sources: (i) the 73 genes recommended by the ACMG for return of incidental findings in clinical exome/genome sequencing^58^; (ii) 13 monogenic diabetes (MODY) genes^59,60^; and (iii) three cardiometabolic genes which have been previously linked to hypertension, blood pressure and obesity^61,62^.

For each gene, we constructed three nested variant burden masks:

1. Pathogenic (ClinVar/ENIGMA) burden: carriers of ≥1 variant annotated as pathogenic in ClinVar^63^ (for BRCA1/2, pathogenic variants from ENIGMA^57^ were used).
2. pLOF burden: carriers meeting criterion (1) or carrying ≥1 LOFTEE^56^ high confidence pLOF variant.
3. Predicted deleterious burden: carriers meeting criterion (1) or (2), or carrying ≥1 missense variant predicted deleterious by ACMG/AMP PP3 evidence (REVEL ≥ 0.932)^64^.

To focus on rare variants, variants with allele frequency > 0.001 were excluded.

### Rare deleterious variants association analyses

For genes with dominant inheritance, dosage was coded as 0, 1, or 2 (with 2 denoting ≥2 qualifying variants). For recessive genes, an indicator was coded 1 for individuals harboring ≥2 qualifying variants in the gene and 0 otherwise. We used linear regression to estimate the association between gene-specific variant dosage and annual healthcare costs, adjusting for age (at end of follow-up), sex, age², age × sex, genotyping array, year of birth, and genetic principal components (PCs 1–20). Analyses were conducted in R v4.2.1. For power, we restricted inference to gene–burden combinations with ≥5 carriers of rare deleterious variants. To account for multiple testing across the three burden masks, we applied a Bonferroni correction within genes (i.e., across the three tests per gene).

### Probability weighted analyses and national extrapolation

To improve representativeness, we performed probability weighted analyses. Weights were derived from estimated UKB participation probabilities based on 14 variables harmonized between UKB and a representative sample of self reported White participants aged 40–69 years from five waves of the Health Survey for England (HSE; n = 22,646)^36^. For national extrapolation, we estimated the size of the UK population aged 40–69 years using data from the Office for National Statistics (https://www.ons.gov.uk/peoplepopulationandcommunity/culturalidentity/ethnicity/articles/ethnicgroupbyageandsexenglandandwales/census2021). We assumed that the frequency of pLOF carriers among UK adults aged 40–69 mirrors that observed in UKB, acknowledging potential underestimation due to healthy volunteer bias^65^.

### Meta analysis of attributable annual inpatient healthcare costs for rare pLOF variants

We performed pLOF burden testing for four significant genes (BRCA1, BRCA2, MSH, APC) in the GEL and GNH cohorts using the same analysis protocol as for UKB. In GNH, covariate adjustment included age (at end of follow up), sex, sex, age2, age*sex, year of birth, and genetic PCs 1-20. Cohort specific estimates were combined using IVW fixed and random effects meta analysis as implemented in the meta package (version 6.2.1) in R.

### SNP validation with FinnGen annotation browser

To assess the novelty of the 380 conditionally-independent SNPs from the primary meta-analyses, the SNPs were tested in the FinnGen annotation browser (https://annopublic.finngen.fi/). This searches for previous significant associations for SNPs in multiple sources including the OpenTargets Genetics platform from 2022 (which includes variants from the GWAS catalogue), FinnGen (Release 12), UK Biobank and Million Veterans Programme meta-analyses, DeCODE pQTLs and more. This provided >50,000 previously discovered SNP-trait associations, with 31 SNPs having no previous associations (**Supplementary Table 17**).

Using LDSC, SNPs in high LD (R2 >= 0.8) with these 31 sentinel SNPs were then extracted from UK Biobank and again searched for previous associations in the FinnGen annotation tool (**Supplementary Table 18**). After this, 14 of the initial 380 SNPs had no associations with the annotation tool and were not in strong LD with SNPs with previous associations.

With these 14 lead SNPs which lacked phenotype annotations, we performed a systematic manual curation using multiple external resources. First, all variants were queried against the December 2025 release of the Open Targets Genetics platform to identify previously reported genome-wide significant associations and fine-mapping evidence. For the remaining SNPs with no genome-wide significant associations in Open Targets, we further screened for sub–genome-wide disease associations using phenome-wide association scans. Specifically, we queried each variant in the FinnGen PheWAS browser (Release R13). We assessed associations with disease endpoints using a nominal significance threshold of P < 0.00016 which is calculated based on the number of disease endpoints tested (PheWAS threshold). Integrating evidence from credible set overlap and phenome-wide association patterns, we then assigned each SNP to predefined interpretative categories capturing likely tagging of known signals, emerging disease associations, and potential novelty (**Supplementary Table 19**).

### Colocalisation analysis using COLOC

To assess the potential shared genetic basis between healthcare-related traits and inpatient cost phenotypes, we performed colocalisation analysis using COLOC. Summary statistics were first selected based on traits included in the INTERVENE consortium study^66^, followed by additional healthcare-related traits hypothesized to influence cost. The traits selected were those for which large GWAS had been conducted and were common diseases or major risk factors. Summary statistics were obtained from multiple sources, including direct downloads from INTERVENE publications, the MVP-UKB-FinnGen meta-analysis database, the GWAS Catalog, PGC, and other consortia such as GIANT (**Supplementary Table 20**). All summary statistics were harmonized into a uniform input format prior to analysis.

Independent sentinel SNPs obtained from the four cost phenotype meta-analyses via GCTA-COJO were used as query SNPs. For each sentinel SNP, a 1 megabase window (500 kb upstream and downstream) was defined, and all overlapping SNPs were included in COLOC testing. Colocalisation analysis was performed using the standard COLOC implementation, modeling quantitative and case-control datasets as appropriate. Posterior probability of shared causal variant (PP4) of ≥0.8 was considered evidence of colocalisation, consistent with recommended COLOC thresholds^67^.

The results of all colocalisation analyses, including trait pairs tested and sentinel SNPs examined, are provided in Supplementary Table COLOC results.

### Polygenic risk score (PGS) weight creation

PGS were created based on each cost phenotype using the summary statistics generated from the primary meta-analyses. These scores were then used in association analyses with both cost and additional phenotypes. In addition to this, a version of each primary meta-analysis was run which excluded either the UK Biobank or FinnGen cohort, so that PGS could be created and tested within these cohorts.

PGS weights were created using LDAK (MegaPRS). For this SNPs were pruned from the meta-analyses to the HapMap3+ SNPs (“HapMap3 plus” set of SNPs defined by Privé et al. specifically for PGS creation^68^).

Polygenic weights were created using the LDAK^69^ software running model bayesr, a power threshold of 0.25 and using a 1000 Genomes reference for the genetic correlations between SNPs. This automatically selected the best model for weights creation for each set of summary statistics. The polygenic weights were then used in the creation of PGS in cohorts of interest using Plink 2.0 –-score methodology.

### PGS evaluation

Polygenic scores (PGS) derived from the full meta-analysis were constructed for all four cost phenotypes and evaluated in two external cohorts: the Netherlands Twin Registry (NTR; N = 15,440) and the China Kadoorie Biobank (CKB; N = 82,389). In each cohort, PGS were generated by applying the corresponding polygenic weights to cohort-specific genotype data following standard quality control and imputation procedures. Scores were calculated as the sum of risk alleles weighted by their effect sizes and were standardized prior to association testing. Associations were evaluated using linear regression with cohort-specific covariates to account for demographic structure and population stratification.

In NTR, each standardized cost PGS was tested for association with five continuous cost outcomes: inpatient + outpatient costs, prescription costs, mental health-related costs, GP costs, and total costs (defined as the sum of inpatient + outpatient, prescription, mental health, and GP costs). Models were adjusted for sex, age at end of follow-up, age², an age-by-sex interaction term, and genetic principal components. In CKB, the inpatient, prescription, and primary care cost PGS were tested against 172 ICD-10 disease outcomes extracted from electronic health records with at least 100 cases, adjusting for region (10 region indicators), sex, age, age², an age-by-sex interaction term, and the first 10 genetic principal components.

For evaluation of the PGS in FinnGen, weights created excluding FinnGen were used in the creation of cost PGS. These PGS were then tested in prediction of the separate cost phenotypes, using the same association model as above.

### Established PGS association with cost phenotypes

Established PGS were generated as described by Kolosov *et al.*^46^ based on the PGS catalog and excluding studies including Finnish cohorts, yielding 3,025 distinct PGS. To minimise overfitting, we excluded PGS identified by the PGS Browser as incorporating FinnGen samples. This classification was based primarily on automated screening of PGS Catalog metadata for the presence of FinnGen among contributing cohorts, followed by manual verification of the highest-performing scores to ensure that no models including FinnGen had been incorrectly labelled as excluding it.

To harmonize trait annotations and group PGS representing similar biological or clinical constructs, each PGS was assigned a canonical trait label using a large language model applied to the original PGS Catalog trait names in batches of 60 scores. Within each canonical trait group, the PGS with the smallest P-value across the univariate analyses was selected as the representative score for downstream analyses, resulting in a final set of 966 PGS.

We then performed univariate association analyses between each standardized PGS and each of the four healthcare cost phenotypes using linear regression, adjusting for previously described covariates: age, sex, an sex*age interaction term, age^2 and the first 10 genetic PCs.

These 966 PGS were subsequently evaluated jointly using multivariable linear regression, in which all PGS were included simultaneously as predictors of each healthcare cost phenotype, together with the same set of covariates.

### Polygenic score construction and parental effect estimation using SNIPAR

To better isolate direct genetic effects, we leveraged parental genetic data in a family-based design within FinnGen. Maternal and paternal PGSs were estimated using the SNIPAR framework in 181,686 parent–offspring trios^43,70^. Of these, 14,210 trios had both parents directly genotyped. For the remaining trios, parental genotypes were imputed using SNIPAR: 60,593 trios had one parent imputed from an offspring–parent duo, and 106,883 trios had both parents imputed using sibling-pair information.

Polygenic scores for inpatient costs were derived using summary statistics from meta-analyses excluding FinnGen and constructed with MegaPRS, as described above. Score calculation and association testing were performed using SNIPAR’s pgs.py utility, incorporating inpatient cost outcomes and covariates from FinnGen.

## Supporting information

GenCost FAQ

Banner Authors

FinnGen Banner Authors

## Acknowledgements and ethical approval

All participants in each contributing cohort provided informed consent for genetic and health-related research in accordance with local regulations. Detailed information on ethical approvals and acknowledgements are provided below for each study.

Padraig Dixon acknowledges the support received from the National Institute for Health and Care Research applied.

## UK Biobank

This research has been conducted using the UK Biobank Resource under Application Number 78537.

## Australian Genetics of Bipolar Disorder Study

### Acknowledgements

We thank the participants for giving their time and support for this project.

### Ethics

Ethics approval for all aspects of the project was obtained from the QIMR Berghofer Human Research Ethics Committee for the GBP study (P3408) and Australian Genetics of Depression Study (P2118). The External Request Evaluation Committee of *Services Australia* approved the GBP study mail-out and the consenting process for linkage to participant Medicare Benefits Schedule (MBS) and Pharmaceutical Benefits Scheme (PBS) data (EREC reference number MI10846 and MI10307, respectively).

### Funding

We acknowledge and thank M. Steffens for her generous donations in loving memory of J. Banks. Data collection was funded and data analysis was supported by the Australian National Health and Medical Research Council (No. APP1138514) to S.E.M.. D.J.S. is supported by a National Health and Medical Research Council Investigator Grant (No. APP1194635). N.G.M. was supported by a National Health and Medical Research Council Investigator Grant (No. APP1172990). S.E.M. is supported by a National Health and Medical Research Council Investigator Grant (No. APP2025674).

## Australian Genetics of Depression Study

### Acknowledgements

We thank all the participants for giving their time to contribute to this study. We wish to thank all the people who helped in the conception, implementation, beta testing, media campaign and data cleaning. We would specifically like to acknowledge Ken Kendler, Patrick Sullivan, Andrew McIntosh and Cathryn Lewis for input on the questionnaire; Lorelle Nunn, Mary Ferguson, Lucy Winkler and Natalie Garden for data and sample collection. Jonathan Davies, Luke Lowrey and Valeriano Antonini for support with IT aspects; Vera Morgan and Ken Kirkby for help with the media campaign.

### Ethics

Ethics approval for all aspects of the project was obtained from the QIMR Berghofer Medical Research Institute Human Research Ethics Committee (P2118). In addition, the AGDS study mail-out and access to participant Medicare Benefits Schedule (MBS) and Pharmaceutical Benefits Scheme (PBS) data were also approved by the *Services Australia* External Request Evaluation Committee (EREC reference number MI3967).

### Funding

The AGDS was primarily funded by National Health and Medical Research Council (NHMRC) of Australia (Grant No. APP1086683) to N.G.M.. This work was further supported by NHMRC grants (No. 1145645, 1078901 and 1087889).

## Estonian Biobank

### Acknowledgements

We want to acknowledge the participants of the Estonian Biobank for their contributions. The Estonian Genome Centre analyses were partially carried out in the High Performance Computing Center, University of Tartu.

### Ethics

The activities of the EstBB are regulated by the Human Genes Research Act, which was adopted in 2000 specifically for the operations of EstBB. Individual level data analysis in EstBB was carried out under ethical approval 1.1-12/624 from the Estonian Committee on Bioethics and Human Research (Estonian Ministry of Social Affairs), using data according to release application 6-7/GI/33520 from the Estonian Biobank.

### Funding

The work of the Estonian Genome Center, University of Tartu was funded by the European Union through Estonian Research Council Grant PRG1291.

### Cohort description

The Estonian Biobank is a volunteer-based biobank with 212,955 participants in the current data freeze^71^. All biobank participants have signed a broad informed consent form and information on ICD-10 codes is obtained via regular linking with the national Health Insurance Fund and other relevant databases, with majority of the electronic health records having been collected since 2004^72^. Analyses were restricted to individuals with European ancestry.

## Genes & Health

### Funding Statement

Genes & Health is/has recently been core-funded by Wellcome (WT102627, WT210561), the Medical Research Council (UK) (M009017, MR/X009777/1, MR/X009920/1), Higher Education Funding Council for England Catalyst, Barts Charity (845/1796), Health Data Research UK (for London substantive site), and research delivery support from the NHS National Institute for Health Research Clinical Research Network (North Thames). We acknowledge the support of the National Institute for Health and Care Research Barts Biomedical Research Centre (NIHR203330); a delivery partnership of Barts Health NHS Trust, Queen Mary University of London, St George’s University Hospitals NHS Foundation Trust and St George’s University of London

Genes & Health is/has recently been funded by Alnylam Pharmaceuticals, Genomics PLC; and a Life Sciences Industry Consortium of AstraZeneca PLC, Bristol-Myers Squibb Company, GlaxoSmithKline Research and Development Limited, Maze Therapeutics Inc, Merck Sharp & Dohme LLC, Novo Nordisk A/S, Pfizer Inc, Takeda Development Centre Americas Inc.

We thank Social Action for Health, Centre of The Cell, members of our Community Advisory Group, and staff who have recruited and collected data from volunteers. We thank the NIHR National Biosample Centre (UK Biocentre), the Social Genetic & Developmental Psychiatry Centre (King’s College London), Wellcome Sanger Institute, and Broad Institute for sample processing, genotyping, sequencing and variant annotation. This work uses data provided by patients and collected by the NHS as part of their care and support. This research utilised Queen Mary University of London’s Apocrita HPC facility, supported by QMUL Research-IT, http://doi.org/10.5281/zenodo.438045

We thank: Barts Health NHS Trust, NHS Clinical Commissioning Groups (City and Hackney, Waltham Forest, Tower Hamlets, Newham, Redbridge, Havering, Barking and Dagenham), East London NHS Foundation Trust, Bradford Teaching Hospitals NHS Foundation Trust, Public Health England (especially David Wyllie), Discovery Data Service/Endeavour Health Charitable Trust (especially David Stables), Voror Health Technologies Ltd (especially Sophie Don), NHS England (for what was NHS Digital) – for GDPR-compliant data sharing backed by individual written informed consent.

Most of all we thank all of the volunteers participating in Genes & Health.

A favourable ethical opinion for the main Genes & Health research study was granted by NRES Committee London – South East (reference 14/LO/1240) on 16 Sept 2014. Queen Mary University of London is the Sponsor, and Data Controller.

## Copenhagen Hospital Biobank

### Acknowledgements

We would like to acknowledge the Novo Nordisk Foundation (grants and NNF14CC0001, NNF17OC0027594, NNF24SA0098829).

## China Kadoorie Biobank

### Acknowledgments

CKB acknowledges the contributions of the study participants, the members of the survey teams in each of the 10 regional centres, and the project development and management teams based at Beijing, Oxford and the 10 regional centres. The CKB baseline survey and the first re-survey were supported by the Kadoorie Charitable Foundation in Hong Kong. The long-term follow-up and subsequent resurveys have been supported by Wellcome grants to Oxford University (212946/Z/18/Z, 202922/Z/16/Z, 104085/Z/14/Z, 088158/Z/09/Z) and grants from the National Natural Science Foundation of China (82192901, 82192904, 82192900) and from the National Key Research and Development Program of China (2016YFC0900500). The UK Medical Research Council (MC_UU_00017/1, MC_UU_12026/2, MC_U137686851), Cancer Research UK (C16077/A29186, C500/A16896) and the British Heart Foundation (CH/1996001/9454), provided core funding to the Clinical Trial Service Unit and Epidemiological Studies Unit at Oxford University for the project. DNA extraction and genotyping were supported by GlaxoSmithKline and the UK Medical Research Council (MC-PC-13049, MC-PC-14135). Computation used the Oxford Biomedical Research Computing (BMRC) facility, a joint development between the Wellcome Centre for Human Genetics and the Big Data Institute supported by Health Data Research UK and the NIHR Oxford Biomedical Research Centre; the views expressed are those of the authors and not necessarily those of the NHS, the NIHR, or the Department of Health.

### CKB Ethics

Ethical approval was obtained from the Ethical Review Committee of the Chinese Centre for Disease Control and Prevention (Beijing, China, 005/2004) and the Oxford Tropical Research Ethics Committee, University of Oxford (UK, 025-04), and all participants provided written informed consent.

## FinnGen

### Ethics statement and materials & methods

Study subjects in FinnGen provided informed consent for biobank research, based on the Finnish Biobank Act. Alternatively, separate research cohorts, collected prior the Finnish Biobank Act came into effect (in September 2013) and start of FinnGen (August 2017), were collected based on study-specific consents and later transferred to the Finnish biobanks after approval by Fimea (Finnish Medicines Agency), the National Supervisory Authority for Welfare and Health. Recruitment protocols followed the biobank protocols approved by Fimea. The Coordinating Ethics Committee of the Hospital District of Helsinki and Uusimaa (HUS) statement number for the FinnGen study is Nr HUS/990/2017.

The FinnGen study is approved by Finnish Institute for Health and Welfare (permit numbers: THL/2031/6.02.00/2017, THL/1101/5.05.00/2017, THL/341/6.02.00/2018, THL/2222/6.02.00/2018, THL/283/6.02.00/2019, THL/1721/5.05.00/2019 and THL/1524/5.05.00/2020), Digital and population data service agency (permit numbers: VRK43431/2017-3, VRK/6909/2018-3, VRK/4415/2019-3), the Social Insurance Institution (permit numbers: KELA 58/522/2017, KELA 131/522/2018, KELA 70/522/2019, KELA 98/522/2019, KELA 134/522/2019, KELA 138/522/2019, KELA 2/522/2020, KELA 16/522/2020), Findata permit numbers THL/2364/14.02/2020, THL/4055/14.06.00/2020, THL/3433/14.06.00/2020, THL/4432/14.06/2020, THL/5189/14.06/2020, THL/5894/14.06.00/2020, THL/6619/14.06.00/2020, THL/209/14.06.00/2021, THL/688/14.06.00/2021, THL/1284/14.06.00/2021, THL/1965/14.06.00/2021, THL/5546/14.02.00/2020, THL/2658/14.06.00/2021, THL/4235/14.06.00/2021, THL/4990/14.02.00/2023 Statistics Finland (permit numbers: TK-53-1041-17 and TK/143/07.03.00/2020 (earlier TK-53-90-20) TK/1735/07.03.00/2021, TK/3112/07.03.00/2021) and Finnish Registry for Kidney Diseases permission/extract from the meeting minutes on 4th July 2019.

The Biobank Access Decisions for FinnGen samples and data utilized in FinnGen Data Freeze 13 include: THL Biobank BB2017_55, BB2017_111, BB2018_19, BB_2018_34, BB_2018_67, BB2018_71, BB2019_7, BB2019_8, BB2019_26, BB2020_1, BB2021_65, BB22-0025-A01, BB22-0025-A03, BB23-0222-A01, BB22-0025-A04, BB22-0025-A06, BB22-0025-A08, THLBB2024_30. Finnish Red Cross Blood Service Biobank 7.12.2017, 13.11.2023, 001-2023, Helsinki Biobank HUS/359/2017, HUS/248/2020, HUS/430/2021 §28, §29, HUS/150/2022 §12, §13, §14, §15, §16, §17, §18, §23, §58, §59, HUS/128/2023 §18, BB22-0025-A01, BB22-0025-A02, BB22-0025-A05, BB22-0025-A07, BB22-0025-A09, BB22-0025-A10, BB22-0025-A03, BB23-0222-A01, BB22-0025-A04, BB22-0025-A06, BB22-0025-A08, Amendment_BB22-0025-A05, Decision allowing to continue data processing until 31st Aug 2027: BB_2021-0140, HUS/150/2022 §12, BB_2021-0139, HUS/150/2022 §13, BB_2021-0161,HUS/150/2022 §14, BB_2021-0164, HUS/150/2022 §15, BB_2021-0169, HUS/150/2022 §16, BB_2021-0170, HUS/150/2022 §17, BB_2021-0179, HUS/150/2022 §18, BB_2022-0262, HUS/150/2022 §58, BB22-0067, HUS/150/2022 §59, Auria Biobank AB17-5154 and amendment #1 (August 17 2020) and amendments BB_2021-0140, BB_2021-0156 (August 26 2021, Feb 2 2022), BB_2021-0169, BB_2021-0179, BB_2021-0161, AB20-5926 and amendment #1 (April 23 2020) and it’s modifications (Sep 22 2021), BB_2022-0262, BB_2022-0256, BB22-0025-A01, BB22-0025-A02, BB22-0025-A03, BB23-0222_A01, BB22-0025-A02, BB22-0025-A05, BB22-0025-A07, BB22-0025-A09, BB22-0025-A10, BB22-0025-A03, BB23-0222-A01, BB22-0025-A04, BB22-0025-A06, BB22-0025-A08, Decision allowing to continue data processing until 31st Aug 2027: AB20-5926, BB_2021-0140, BB_2021-0156, BB_2021-0161, BB_2021-0161, BB_2021-0164, BB_2021-0169, BB_2021-0179, BB_2022-0262, Biobank Borealis of Northern Finland_2017_1013, 2021_5010, 2021_5010 Amendment, 2021_5018, 2021_5018 Amendment, 2021_5015, 2021_5015 Amendment, 2021_5015 Amendment_2, 2021_5023, 2021_5023 Amendment, 2021_5023 Amendment_2, 2021_5017, 2021_5017 Amendment, 2022_6001, 2022_6001 Amendment, 2022_6006 Amendment, 2022_6006 Amendment_2, BB22-0067, 2022_0262, 2022_0262 Amendment, BB22-0025-A01, BB22-0025-A02, BB22-0025-A05, BB22-0025-A07, BB22-0025-A09, BB22-0025-A10, BB22-0025-A03, BB23-0222-A01, BB22-0025-A04, BB22-0025-A06, BB22-0025-A08, Decision allowing to continue data processing until 31st Aug 2027: BB/2021/5015, BB/2021/5017, BB/2021/5018, BB/2021/5023, BB/2022/6006, BB/2022/6001, BB/2022-0262, BB/2021/5010, Biobank of Eastern Finland 1186/2018 and amendment 22§/2020, 53§/2021, 13§/2022, 14§/2022, 15§/2022, 27§/2022, 28§/2022, 29§/2022, 33§/2022, 35§/2022, 36§/2022, 37§/2022, 39§/2022, 7§/2023, 32§/2023, 33§/2023, 34§/2023, 35§/2023, 36§/2023, 37§/2023, 38§/2023, 39§/2023, 40§/2023, 41§/2023, BB22-0025-A01, BB22-0025-A02, BB22-0025-A05, BB22-0025-A07, BB22-0025-A09, BB22-0025-A10, BB22-0025-A03, BB23-0222-A01, BB22-0025-A04, BB22-0025-A06, BB22-0025-A08, Decision allowing to continue data processing until 31st Aug 2027: MO-BB_2021-0179-A0, MO-BB_2021-0156_PRE-A01, BB_2021-0140, MO-BB_2021-0170_PRE-A0, MO-BB_2021-0169-A01, MO-BB_2022-0256-A01, MO-BB_2021-0161-A01, MO-BB_2021-0161-A02, BB22-0067-A01, MO-BB_2022-0262-A0, Finnish Clinical Biobank Tampere MH0004 and amendments (21.02.2020 & 06.10.2020), BB2021-0140 8§/2021, 9§/2021, §9/2022, §10/2022, §12/2022, 13§/2022, §20/2022, §21/2022, §22/2022, §23/2022, 28§/2022, 29§/2022, 30§/2022, 31§/2022, 32§/2022, 38§/2022, 40§/2022, 42§/2022, 1§/2023, BB2021-0140, BB22-0025-A01, BB_2021-0161, BB22-0025-A02, BB22-0025-A05, BB22-0025-A07, BB22-0025-A09, BB22-0025-A10, BB22-0025-A03, BB23-0222-A01, BB22-0025-A04, BB22-0025-A06, BB22-0025-A08, Decision allowing to continue data processing until 31st Aug 2027: BB_2021-0140, BB_ 2021-0161, BB_ 2021-0179, BB_ 2021-0156, BB_ 2021-0169, BB_ 2021-0170, BB22-0067-A01, Central Finland Biobank 1-2017, BB_2021-0169, BB_2021-0179, BB_2022-0256, BB_2022-0262, Decision allowing to continue data processing until 31st Aug 2027 for projects: BB_2021-0179, BB22-0067,BB_2022-0262, BB_2021-0170, BB_2021-0164, BB_2021-0161, and BB_2021-0169, BB22-0025-A01, BB22-0025-A02, BB22-0025-A05, BB22-0025-A07, BB22-0025-A09, BB22-0025-A10, BB22-0025-A03, BB23-0222-A01, BB22-0025-A04, BB22-0025-A06, BB22-0025-A08, Terveystalo Biobank STB 2018001 and amendment 25th Aug 2020, Finnish Hematological Registry and Clinical Biobank decision 18th June 2021, Amendment 2nd January 2024 and Arctic biobank P0844: ARC_2021_1001, ARC_2023_3003 (BB22-0025-A01), BB22-0025-A03, BB23-0222-A01, BB22-0025-A04, BB22-0025-A06, BB22-0025-A08.

### Acknowledgements

We want to acknowledge the participants and investigators of the FinnGen study. The FinnGen project is funded by two grants from Business Finland (HUS 4685/31/2016 and UH 4386/31/2016) and the following industry partners: AbbVie Inc., Alnylam Pharmaceuticals, Inc., AstraZeneca UK Ltd, Bayer AG, Biogen MA Inc., Boehringer Ingelheim International GmbH, Bristol Myers Squibb Inc. (and Celgene Corporation & Celgene International II Sàrl), Genentech Inc., GlaxoSmithKline Intellectual Property Development Ltd., Johnson&Johnson Innovative Medicine Inc., Maze Therapeutics Inc., Merck Sharp & Dohme LCC, Novartis AG, Pfizer Inc. and Sanofi US Services Inc. Following biobanks are acknowledged for delivering biobank samples to FinnGen: Auria Biobank (www.auria.fi/biopankki), THL Biobank (www.thl.fi/biobank), Helsinki Biobank (www.helsinginbiopankki.fi), Biobank Borealis of Northern Finland (https://www.ppshp.fi/Tutkimus-ja-opetus/Biopankki/Pages/Biobank-Borealis-briefly-in-English.aspx), Finnish Clinical Biobank Tampere (www.tays.fi/en-US/Research_and_development/Finnish_Clinical_Biobank_Tampere), Biobank of Eastern Finland (www.ita-suomenbiopankki.fi/en), Central Finland Biobank (www.ksshp.fi/fi-FI/Potilaalle/Biopankki), Finnish Red Cross Blood Service Biobank (www.veripalvelu.fi/verenluovutus/biopankkitoiminta), Terveystalo Biobank (www.terveystalo.com/fi/Yritystietoa/Terveystalo-Biopankki/Biopankki/) and Arctic Biobank (https://www.oulu.fi/en/university/faculties-and-units/faculty-medicine/northern-finland-birth-cohorts-and-arctic-biobank). All Finnish Biobanks are members of BBMRI.fi infrastructure (https://www.bbmri-eric.eu/national-nodes/finland/). Finnish Biobank Cooperative –FINBB (https://finbb.fi/) is the coordinator of BBMRI-ERIC operations in Finland. The Finnish biobank data can be accessed through the Fingenious® services (https://site.fingenious.fi/en/) managed by FINBB.

## Generation Scotland

### Ethics

All components of Generation Scotland received ethical approval from the NHS Tayside Committee on Medical Research Ethics (REC Reference Number: 05/S1401/89). All participants provided broad and enduring written informed consent for biomedical research. Generation Scotland has also been granted Research Tissue Bank status by the East of Scotland Research Ethics Service (REC Reference Number: 15/0040/ES), providing generic ethical approval for a wide range of uses within medical research. This study was performed in accordance with the Helsinki declaration.

## Funding/Acknowledgements

This research was funded in whole, or in part, by the Wellcome Trust (104036/Z/14/Z). Generation Scotland received core support from the Chief Scientist Office of the Scottish Government Health Directorates (CZD/16/6) and the Scottish Funding Council (HR03006). DNA profiling of the Generation Scotland samples was carried out by the Genetics Core Laboratory at the Edinburgh Clinical Research Facility, Edinburgh, Scotland, and was funded by the Medical Research Council UK.

## Genomics England

### Acknowledgements

We gratefully acknowledge the participants of the National Genomic Research Library (NGRL), whose contributions made this research possible. Secure access to the NGRL under project ID 883 was provided by Genomics England, which delivers the NGRL in partnership with NHS England, and is wholly owned by the UK Department of Health and Social Care. The NGRL contains participants’ health data collected by the NHS as part of their care, along with samples and data from their participation in research, for which fully informed consent has been obtained. This includes genomic and clinical data provided through the NHS Genomic Medicine Service, as well as data obtained through research studies, including the 100,000 Genomes Project and the Generation Study, both of which are delivered in partnership with the NHS, and from other research cohorts involving external collaborators.

## Data access

Data from the National Genomic Research Library (NGRL)^73^ used in this research are available within the secure Genomics England Research Environment. Access to NGRL data is restricted to adhere to consent requirements and protect participant privacy.

Data used in this research include derived phenotype files constructed for genome-wide association analyses using NHS healthcare cost data linked to the Genomics England Main Programme v8 (2019-11-28), restricted to affected probands and relatives. Healthcare costs were estimated from Hospital Episode Statistics (HES) inpatient records using the 2018/19 NHS Reference Cost Grouper and National Reference Cost Schedule. Additional datasets from Main Programme v9 used in phenotype construction included participant_summary, death details, aggregate_gVCF_sample_stats, and rare_diseases_pedigree_member. Derived phenotype files are stored within the Research Environment at: /re_gecip/health_economics/kzguro/gwas_costs/phenotypes/.

Genotype data were obtained from the Genomics England Main Programme aggregate gVCF dataset (aggV2, Strelka pipeline), accessed via pre-filtered BGEN files. High-quality linkage disequilibrium-pruned SNP sets were used for ancestry and relatedness adjustment, and unrelated individuals were identified using precomputed relatedness metrics provided within the Research Environment. GWAS summary statistics generated in this study are stored at: /re_gecip/health_economics/kzguro/gwas_costs/phenotypes/inpatients/EUR/.

For whole-exome sequencing (WES) analyses, genotype data were accessed as PLINK2 pgen files with low-quality genotypes masked. Variants were annotated using Variant Effect Predictor (VEP) version 109. Derived WES analysis files are stored at: /re_gecip/health_economics/kzguro/gencost_wes/.

Access to NGRL data is provided to approved researchers who are members of the Genomics England Research Network, subject to institutional access agreements and research project approval under participant-led governance. For more information on data access, visit: https://www.genomicsengland.co.uk/research

## Data availability

Summary statistics of the primary meta-analyses and stratified meta-analyses will be available through the European Bioinformatics Institute GWAS Catalog (https://www.ebi.ac.uk/gwas/) upon submission.

PGS-weights derived using MegaPRS will likewise be made publicly available through the PGS-catalog.

## Code availability

All code is available at: https://github.com/Sabor117/GenCost

