## Supplementary Information for "Quantifying the contribution of genetic variation to healthcare expenditure across diverse healthcare systems"

**Multinational genomewide association study identifies genetic variants associated with healthcare costs: the GenCost consortium**

**Supplementary information - Cost phenotype definitions**

**UK Biobank (UKBB)**

Inpatient costs were derived from UK Biobank NHS hospital record tables (UK Biobank field 41259, HESIN data), primary care costs derived from GP visits (field 42040, GP clinical events) and prescriptions from GP prescriptions (field 42039, prescription records).

All records of each type were restricted at absolute maximum to the period between 01/06/2006 (a cut-off used previously for hospital records to ensure data integrity) and 19/12/2022 (the final date in the UK Biobank death register at time of conducting this analysis). For inpatient hospital records, the period of follow-up for each person was then restricted to the dates between their entry into UK Biobank (between 2006 and 2010) and *either* 31/10/2021 (one year prior to the final hospital record in the data) *or* their date of death. For both prescriptions and GP data follow-up was defined as being either the earliest GP visit/prescription date or entry into UK Biobank and then calculated until 31/07/2017 and 30/09/2017 respectively (the final dates in the prescription and GP datasets) or the date of death of the individual.

For inpatient data, each hospital record in the defined period of follow-up was input into the NHS Casemix Grouper software (<https://digital.nhs.uk/services/national-casemix-office/downloads-groupers-and-tools>) this includes diagnoses, days in hospital and associated admin codes. This tool outputs a hospital related group (HRG) code for each input hospital episode, and each code can be associated with a mean cost from the 2022 NHS National Schedule reimbursement table. Data for this table was modulated to create proportional mean costs for every HRG across multiple scenarios (elective care, non-elective, day-care, and more, whilst also excluding some such as paediatric care).

For prescriptions, UK Biobank prescriptions from GPs were present for ~45% of the cohort. Due to inconsistent naming of drugs and appliances prescribed, a custom table of prescriptions and costs based on the 2022 NHS Drug Tariff was created in order to assign costs to these prescriptions and applications, achieving 95% coverage of the prescriptions made in the period of follow-up of the analysis.

For primary care costs, the average cost of GP visits in the UK in 2022 (£42, as per <https://www.england.nhs.uk/east-of-england/2023/12/08/quarter-of-a-million-more-seen-by-gps-in-the-east-of-england-during-october-as-costs-of-no-shows-revealed/>) was multiplied per number of visits for each individual in the period of follow-up.

For each cost phenotype, a proportion of individuals were also defined as having a healthcare cost of £0, I.e. healthy individuals. As hospital records were available for every individual in UK Biobank, individuals without hospital records were kept as zero-cost individuals. However, as the NHS Grouper failed in producing HRGs in a small proportion (<1%) of episodes, any individual who only had failed HRG episodes in the period of follow-up were excluded from the analysis as these may not have been true zeros. As the GP and prescription data is only present for a subset of the cohort, the definition of true zeroes was different. For these phenotypes, an individual was only considered to be zero-cost if they were present within the dataset overall (e.g. had a prescription or GP visit prior to the follow-up cut-off) but did not have any visits or prescriptions recorded during the period of follow-up.

With the total overall cost created for each of the three phenotypes, the costs were then converted into annual prices (divide total cost by number of days in follow-up period, divided by 365.25), then from 2022 prices to 2019 prices using the Bank of England consumer price index calculator (<https://www.bankofengland.co.uk/monetary-policy/inflation/inflation-calculator>, £1 in 2022 = £0.89 in 2019), and then from pounds into euros using the average exchange rate of 2019 (£1 = €1.1405) from<https://www.exchangerates.org.uk/GBP-EUR-spot-exchange-rates-history-2019.html>. As these corrected annual costs would then be natural-log transformed for GWA analysis, all costs of <€1 were lastly converted into €1 for the log-transformation.

**Genes and Health (G&H)**

The procedure for the G&H cohort followed very closely the protocol for UK Biobank. Inpatient data stemmed from hospital records subjected to Casemix Grouper processing (using the Linux-specific AutoGrouper software), and prescription drug costs were processed using the same custom drug tables created for UK Biobank. Following the same procedure for UK Biobank these costs were thus also corrected to 2019 prices in Euros utilising the same values and schema.

Follow-up for inpatient costs was defined using fixed calendar cut-offs aligned to the UK Biobank approach, spanning 01/06/2006 to 31/12/2022 (the latest date available in the death register at the time of analysis). For prescription costs the final cut-off date was extended to 31/12/2023.

For inpatient costs, individual follow-up was calculated from 01/06/2006 until 31/12/2022 or date of death, whichever occurred first. For prescription costs, follow-up accrued from 01/06/2006 or the date of the earliest recorded prescription (if later than 01/06/2006) until the date of their final recorded prescription. Individuals with no hospital utilisation records in G&H were assigned zero inpatient cost, consistent with UK Biobank practice. Also in accordance with UK Biobank, the prescriptions analysis was restricted to individuals with at least one recorded prescription, and therefore did not include true zero-cost observations.

**Genomics England (GE)**

Healthcare cost phenotypes for Genomics England (GEL) participants were derived from Hospital Episode Statistics (HES) data using the NHS Reference Cost Grouper (2018/19) and the corresponding National Reference Cost Schedule, with all costs expressed in 2019 GBP values (2018/19 National Cost Collection). All available HES records for the full GEL cohort (probands and relatives) were initially costed to create a harmonised, reusable dataset. For the present analysis, a subset restricted to affected probands and relatives from data release *main_programme_v8_2019_11-28* was used. Inpatient activity was grouped using the NHS Grouper and assigned unit costs according to the national reimbursement schedule. No additional consumer price index adjustment was required, as costs were already standardized to 2019 prices at source. Prices were converted to Euros using the same exchange rate as UK Biobank.

Follow-up was defined at the individual level. For each participant, the start of follow-up corresponded to the first recorded interaction with the English NHS (including birth episodes where applicable), and the end of follow-up was defined as the last available HES record prior to the November 2019 data release. In downstream analyses, age at end of follow-up was defined as age at the HES end date (epiend) or age at death if death occurred earlier. Individuals without any HES records were not included in the costed dataset provided for analysis; absence of records may reflect care received outside NHS England rather than true zero healthcare utilisation. Consequently, “zero cost” status was interpreted only among individuals with available HES data within the defined observation window.

**Estonian Biobank (EstBB)**

EstBB is a volunteer-based biobank with 212,955 participants in the current data freeze [PMID:40188112]. All biobank participants have signed a broad informed consent form and information on ICD-10 codes is obtained via regular linking with the national Health Insurance Fund and other relevant databases, with majority of the electronic health records having been collected since 2004 [PMID:24518929]. Analyses here were limited to participants of European ancestry (n = 205,203 for this analysis).

**a. Follow-up period:**

Follow-up time for each individual was defined as the period from 01/01/2005 until the date of death or 31/12/2019, whichever came first. These dates were chosen to ensure data integrity by avoiding irregularities caused by the COVID-19 pandemic and leveraging a period of relatively stable inflation in Estonia.

**b. Cost origin and c. Cost calculation method:**

Healthcare cost data for EstBB participants were derived from the Estonian Health Insurance Fund (EHIF) billing database. EHIF costs are regulated nationally by law under the “Tervisekassa tervishoiuteenuste loetelu” (EHIF Service List - https://www.riigiteataja.ee/akt/128062019011), which defines fixed reimbursement prices and conditions for each service. These prices are not negotiable and apply uniformly across Estonia. This legal framework ensures that cost values are standardized and reflect real reimbursement expenses. For example, general practitioners are reimbursed using a capitation-based formula, with monthly rates defined by patient age group and practice location. Inpatient and outpatient costs are similarly priced per diagnosis-related service or procedure code, as outlined in the official reimbursement regulation.

Records included inpatient hospital stays, outpatient specialist visits, primary care (general practitioner) visits, and prescription reimbursements. Only records clearly attributable to one of these four categories were included. Other cost types (e.g. laboratory, rehabilitation, transportation) were excluded due to ambiguity in classification. For each individual, total costs per year were calculated separately for inpatient, outpatient, primary care, and prescription records, based on EHIF reimbursement amounts.

**d. Zero-cost individuals:**

Individuals with no records in a given category during their follow-up were treated as having zero cost for that category.

**e. Inflation correction:**

Cost data were aggregated into annual person-level totals, then adjusted to 2019 euros using Estonian consumer price index data provided by Statistics Estonia (via Andres Võrk; https://www.stat.ee/en/find-statistics/statistics-theme/finance/prices/consumer-price-index).

The final dataset included approximately 14.1 million general practitioner visit entries, 10.8 million outpatient specialist visits, 618 thousand inpatient stays, and 19.7 million prescription records. Log-transformation (natural log) of annual costs was performed at the phenotype preparation stage. Individuals with €0 annual cost were excluded from the GWAS analyses to enable transformation.

The activities of the EstBB are regulated by the Human Genes Research Act, which was adopted in 2000 specifically for the operations of EstBB. Individual-level data analysis in EstBB was carried out under ethical approval 1.1-12/624 from the Estonian Committee on Bioethics and Human Research (Estonian Ministry of Social Affairs), using data according to release application 6-7/GI/33520 from the Estonian Biobank.

**The Australian Genetics of Depression Study (AGDS) and Australian Genetics of Bipolar Disorder Study (GBP)**

Primary care costs in each cohort were calculated from consented linkage of Medicare Benefits Schedule (MBS) records. Drug costs in each cohort were calculated from consented linkage of Pharmaceutical Benefits Scheme (PBS) records. The data window for the AGDS cohort was 1/07/2013-31/12/2017 and 30/06/2014-30/06/2019 for the GBP cohort.

Primary care costs were estimated from MBS records after services not charged by general practitioners (GPs) were excluded. The fee listed in the Medicare Benefits Schedule ("Schedule Fee") charged for each service were summed across each calendar year. The total costs in each year were then inflation-adjusted using consumer price index (CPI) derived inflation rates, where the index reference base for the CPI rates was 2011-12, and expressed in 2019 Australian dollar prices. Annual total costs were then calculated by dividing the total costs summed across all years by the number of years the MBS data were available for each participant. Participants who did not have any MBS records in the data window were not included in the analyses. Fewer than 10 individuals were present in the cohorts with no records and were not included as zero costs.

The cost for each drug in the PBS records was calculated as the sum of the Net Benefit fee (the benefit that the Australian Government paid to the Pharmacy) and Patient Contribution fee (the fee paid by the patient). The dollar amount for each dispensed drug was summed across each calendar year. The total costs in each year were then inflation-adjusted using CPI derived inflation rates, where the index reference base for the CPI rates was 2011-12, and expressed in 2019 Australian dollar prices. Annual total costs were then calculated by dividing the total costs summed across all years by the number of years the PBS data were available for each participant. Participants who did not have any PBS records in the data window were not included in the analyses.

**Mass General Brigham Biobank (MGBB)**

Inpatient costs were derived from electronic health records of encounters, diagnoses, and procedures. Diagnoses and procedures coded with International Classification of Diseases Version 9 (ICD-9) and ICD-10. Each encounter, consisting of a set of diagnoses and procedures, were converted to diagnosis-related groups (DRGs) using the Medicare Severity-Diagnosis Related Group conversion software in Java from the Centers for Medicare and Medicaid Services. Each DRG was converted to costs using Medicare’s standardized national costs derived for inpatient hospitals. The follow-up period was captured between January 1, 2005 and April 6, 2022. Annual costs were calculated by dividing each cost by the amount of follow-up time. There were no individuals with missing costs, as only inpatient encounters were considered and all inpatient encounters had associated costs. The costs were expressed in 2021 prices in dollars and were not adjusted to 2019 prices via any CPI.

**Copenhagen Hospital Biobank (CHB)**

Inpatient and outpatient healthcare costs were derived from the Danish National Health Registry, using the Diagnosis-Related Group (DRG) subset. For each hospital contact occurring during the follow-up period, costs were assigned based on the corresponding DRG code. Cost estimates reflected year-specific reimbursement fees applicable at the time of the hospital episode. Follow-up for inpatient and outpatient hospital data was restricted to the period between 2005, corresponding to the registry data integrity cut-off, and 2017, the final available record date, or until the date of death if this occurred earlier.

Prescription drug costs were obtained from the Danish National Medication Registry. Each dispensed prescription was assigned a cost based on its associated VNR code, with prices varying by calendar year to reflect changes in medication pricing over time.

All healthcare costs were aggregated at the individual level, annualized, and standardized to 2019 monetary values. Costs were converted to Euros using the average exchange rate for 2019, consistent with the approach applied in the UK Biobank analyses. Due to the clinical characteristics of the Copenhagen Hospital Biobank cohort, zero-cost individuals were not observed. Participants in this biobank are generally relatively ill, as inclusion requires blood sampling for clinical purposes such as blood typing and anticipated surgical procedures with potential transfusion needs. Consequently, unlike population-based cohorts such as the UK Biobank, this dataset does not include partially healthy individuals with no recorded healthcare expenditures.

**Generation Scotland (GS20K)**

Inpatient healthcare costs for Generation Scotland were derived from electronically linked NHS hospital records. Hospital episodes occurring prior to 2000 were excluded from the analysis. Follow-up was defined from cohort entry until 30 April 2022, corresponding to the most recent date of available linked health records at the time of analysis, or until date of death if earlier, yielding a maximum follow-up duration of 22 years and 4 months. Recorded hospital episodes were processed using the NHS Hospital Resource Group (HRG) Grouper software to assign HRG codes (<https://digital.nhs.uk/services/national-casemix-office/downloads-groupers-and-tools/hrg4-2023-24-consultation-grouper>). These HRG codes were subsequently mapped to costs in British Pounds Sterling using the NHS National Tariff Payment System (NTPS) reference tables (<https://www.england.nhs.uk/wp-content/uploads/2021/02/20-21NT_Annex_A_National_tariff_workbook.xlsx>). To account for temporal changes in healthcare costs, all monetary values were adjusted for inflation using the UK Government CPI indices, with 2019 specified as the reference year (<https://www.ons.gov.uk/file?uri=/economy/inflationandpriceindices/datasets/consumerpriceinflation/current/consumerpriceinflationdetailedreferencetables.xlsx>). Individuals with available linked health records but no recorded inpatient episodes during follow-up were retained in the analysis and assigned zero cost, representing healthcare non-utilizers or relatively healthy individuals.

**FinnGen**

Costs were derived from several national registries. Medication costs were derived from the KELA registry, inpatient costs were derived from the HILMO registry, outpatient costs were derived from both the HILMO and AVOHILMO registry. Medication costs were derived from average costs associated with each drug product number capturing both count and dose. Inpatient costs were calculated by adding the fixed costs associated with each admission and the costs per day multiplied by the length of stay. Costs from the KELA and HILMO registries were considered between 2000 and 2020, while costs from the AVOHILMO registry was considered between 2011 and 2020 to account for the latter initiation of the AVOHILMO registry. Annual costs were calculated by dividing each cost by the amount of follow-up time in each registry. Individuals without records in the AVOHILMO registry were considered to be users of private insurance and were assigned the median costs. Other missing costs were assigned zero values. Costs were expressed in 2017 prices and were not converted to 2019 prices via CPI.

**Qatar Genome Program (QGP)**

Healthcare costs were derived from electronic health record (EHR)–recorded hospital utilisation between 2014 and 2023 using an average unit-cost approach. For each individual, total cost was calculated as the sum across encounter types. Inpatient cost was estimated by multiplying the number of inpatient days (length of stay) by an average cost per inpatient day, and outpatient cost by multiplying the number of outpatient visits by an average cost per outpatient visit. The combined inpatient + outpatient phenotype was defined as the sum of these inpatient and outpatient components. Individuals with no recorded EHR encounters during 2014–2023 were assigned zero cost. For the GenCost phenotype, annual cost was estimated by dividing total accumulated cost by each individual’s observed follow-up time, yielding a cost per person-year.

**Netherlands Twin Registry (NTR)**

Healthcare costs in the NTR cohort were ascertained over the period 2017 to 2022, with follow-up defined for each individual from 1 January 2017 until 31 December 2022 or until they were no longer registered as a Dutch citizen, for example due to emigration or death. All costs were derived directly from amounts billed to Dutch health insurance companies, through which essentially all healthcare costs in the Netherlands are processed. No additional reimbursement tools, grouping software, or external cost tables were required to assign costs. Thus, the cost phenotypes reflect observed insurance-billed healthcare expenditure during the defined follow-up period.

All zero-cost individuals were considered true zeroes, as nearly all registered Dutch citizens residing in the Netherlands are covered by healthcare insurance, aside from rare exceptions such as abstention for religious reasons. Accordingly, the absence of billed healthcare costs during follow-up was interpreted as absence of healthcare expenditure rather than missing data. Individuals with missing non-zero cost data were assumed to have likely emigrated or died during follow-up. To harmonize costs across analyses, all costs were converted to 2019 prices using consumer price indices obtained from Statistics Netherlands (CBS, <https://www.cbs.nl/nl-nl/cijfers/detail/83131NED>).
