## Supplementary Figures for "Quantifying the contribution of genetic variation to healthcare expenditure across diverse healthcare systems"

**Supplementary Figure 1.** Histograms with density overlays for cost phenotypes (blue = inpatient costs, orange = prescription costs, purple = inpatient + outpatient costs, green = primary care costs) of the following cohorts:

**A - Australian Genetics of Depression Study (AGDS)**

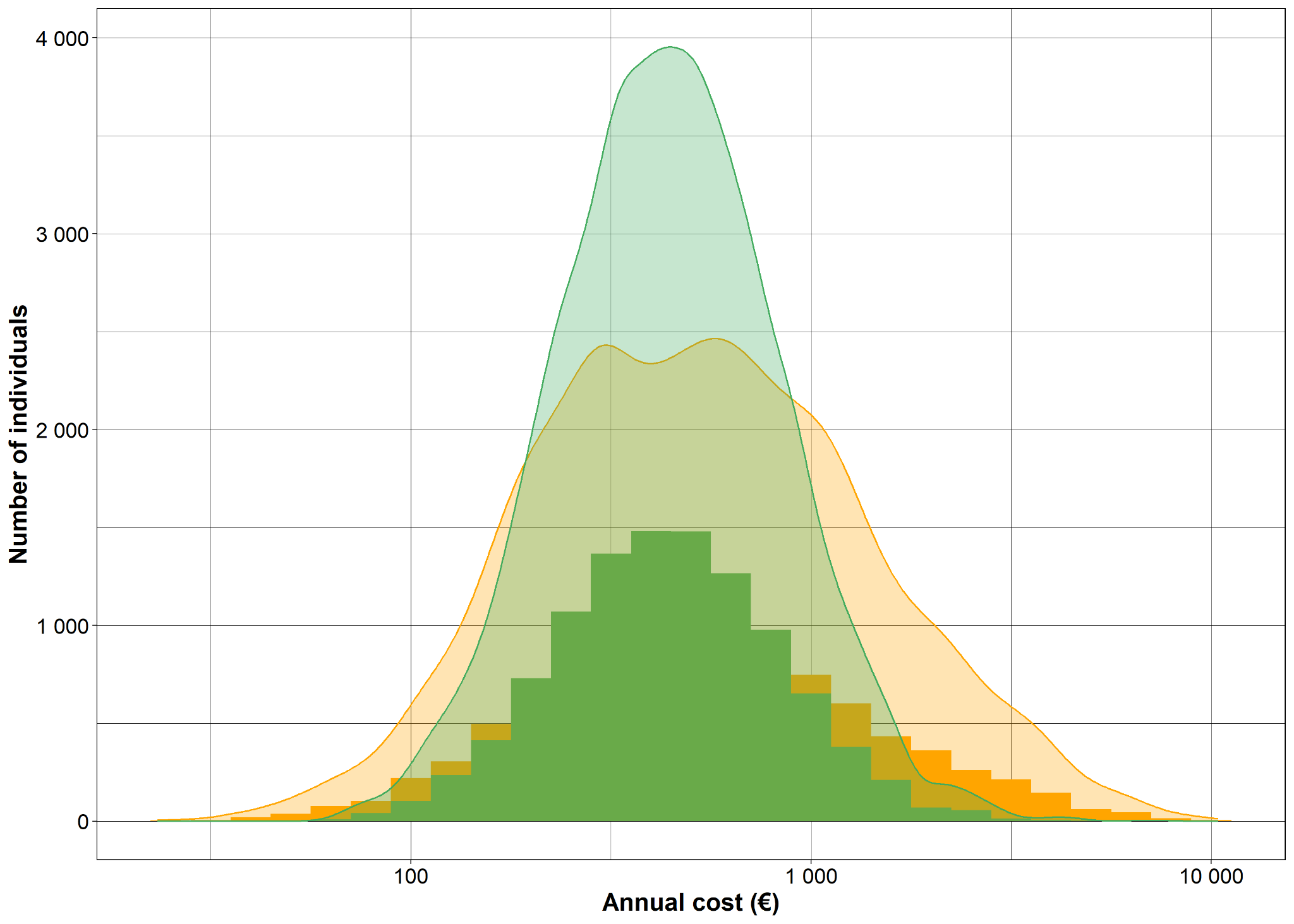

**B - Australian Genetics of Bipolar Disorder (GBP)**

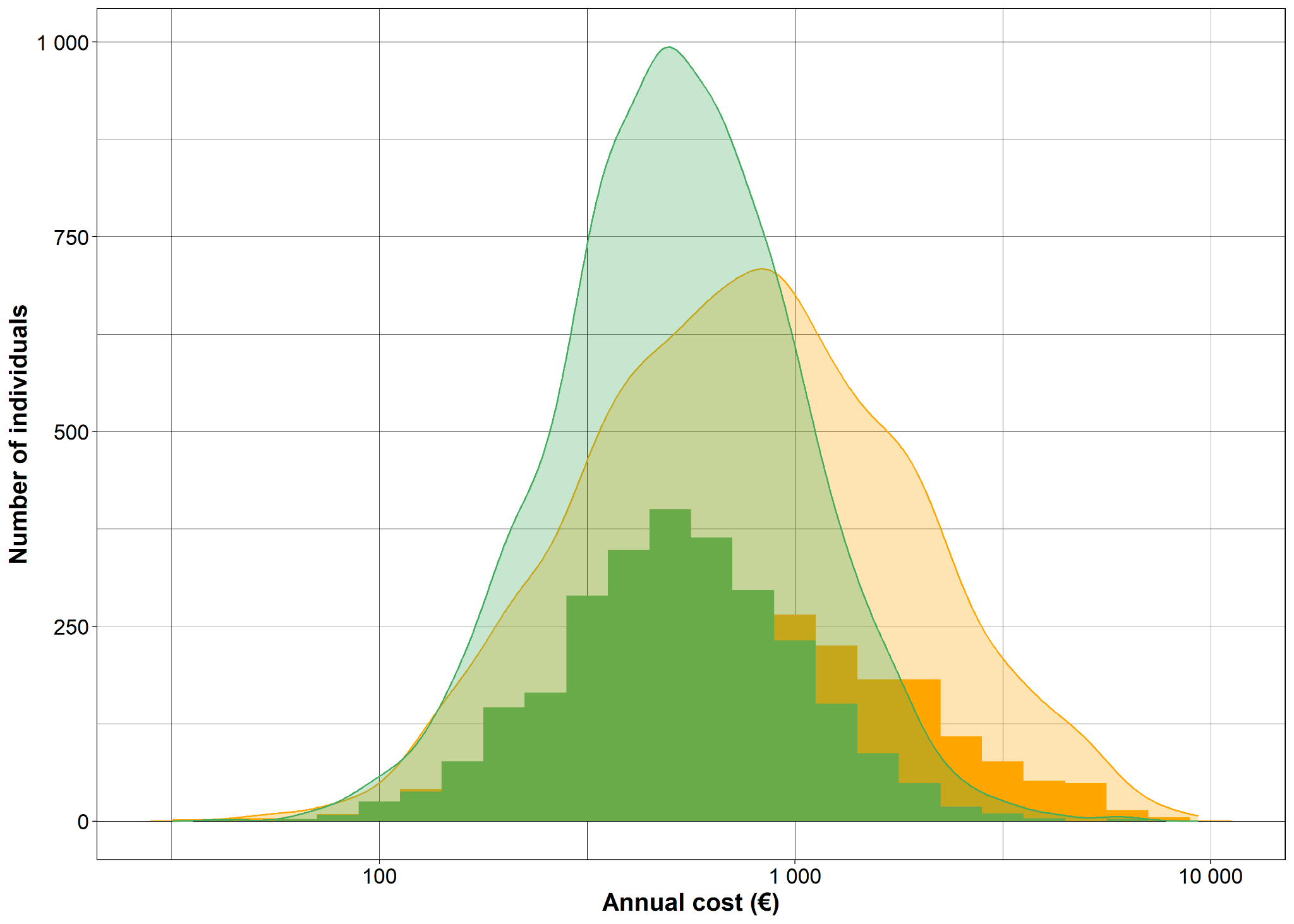

**C - Copenhagen Hospital Biobank (CHB)**

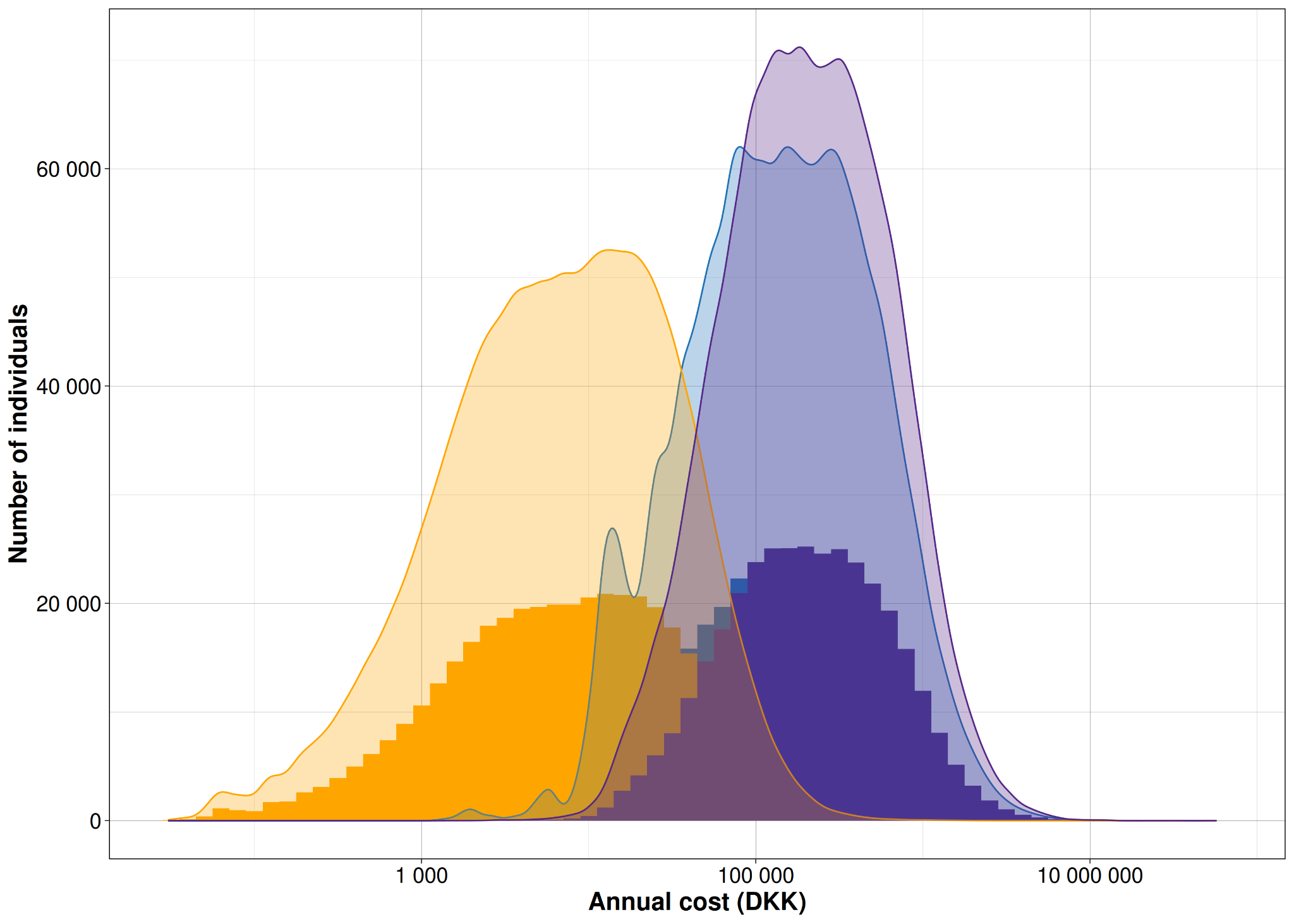

**D - Estonian Biobank (EstBB)**

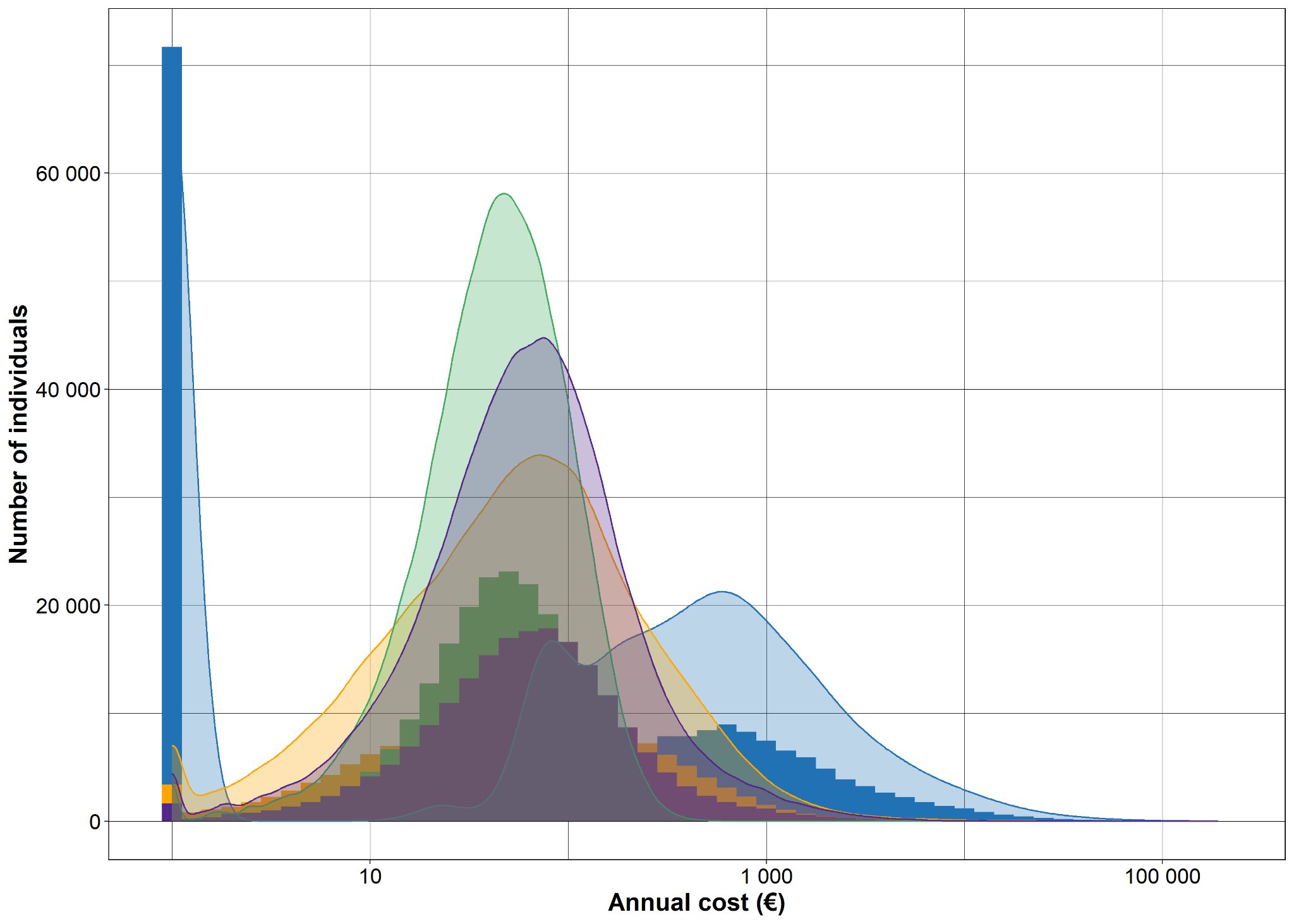

**E - Generation Scotland (GS)**

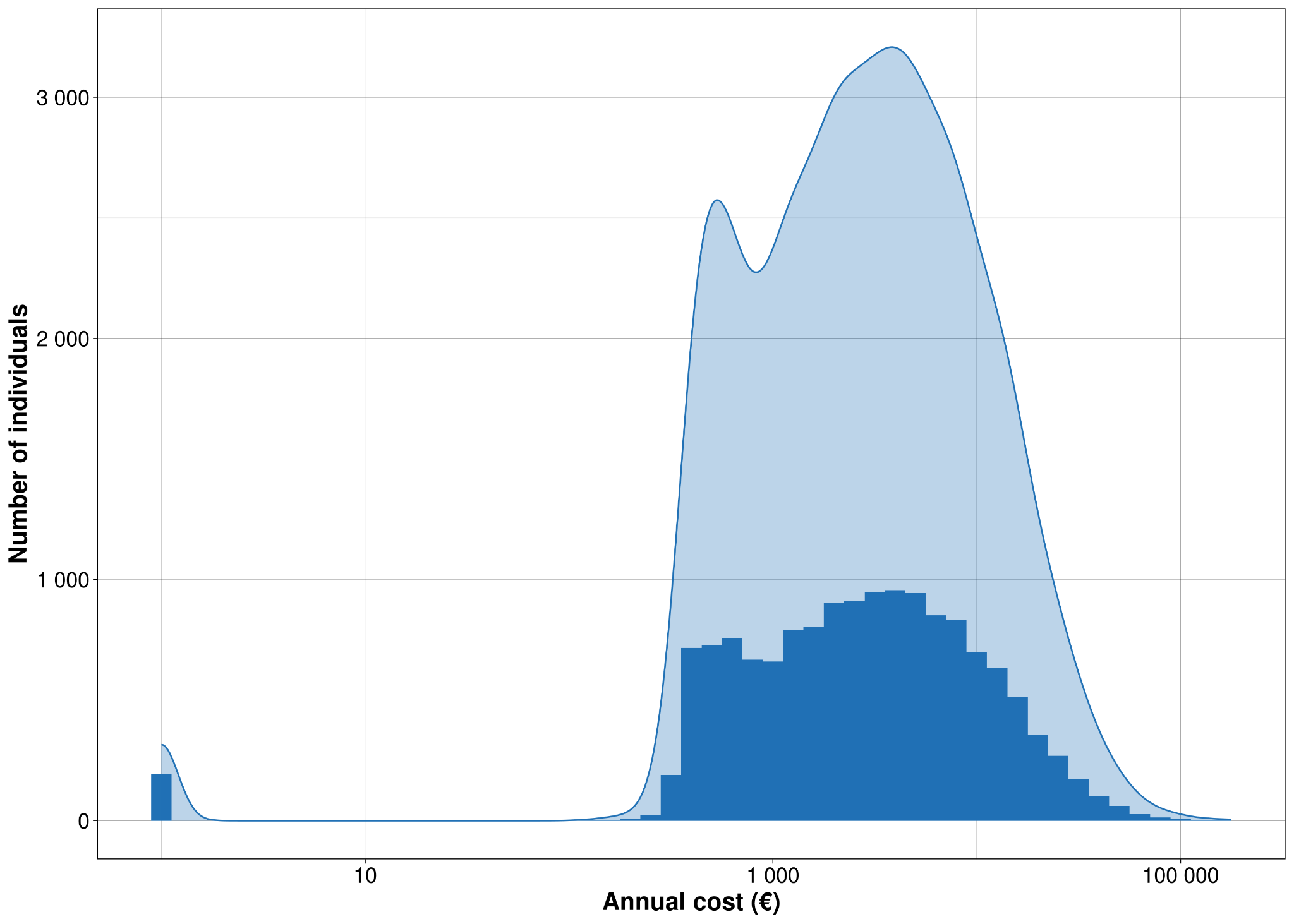

**F - Genomics England (GE)**

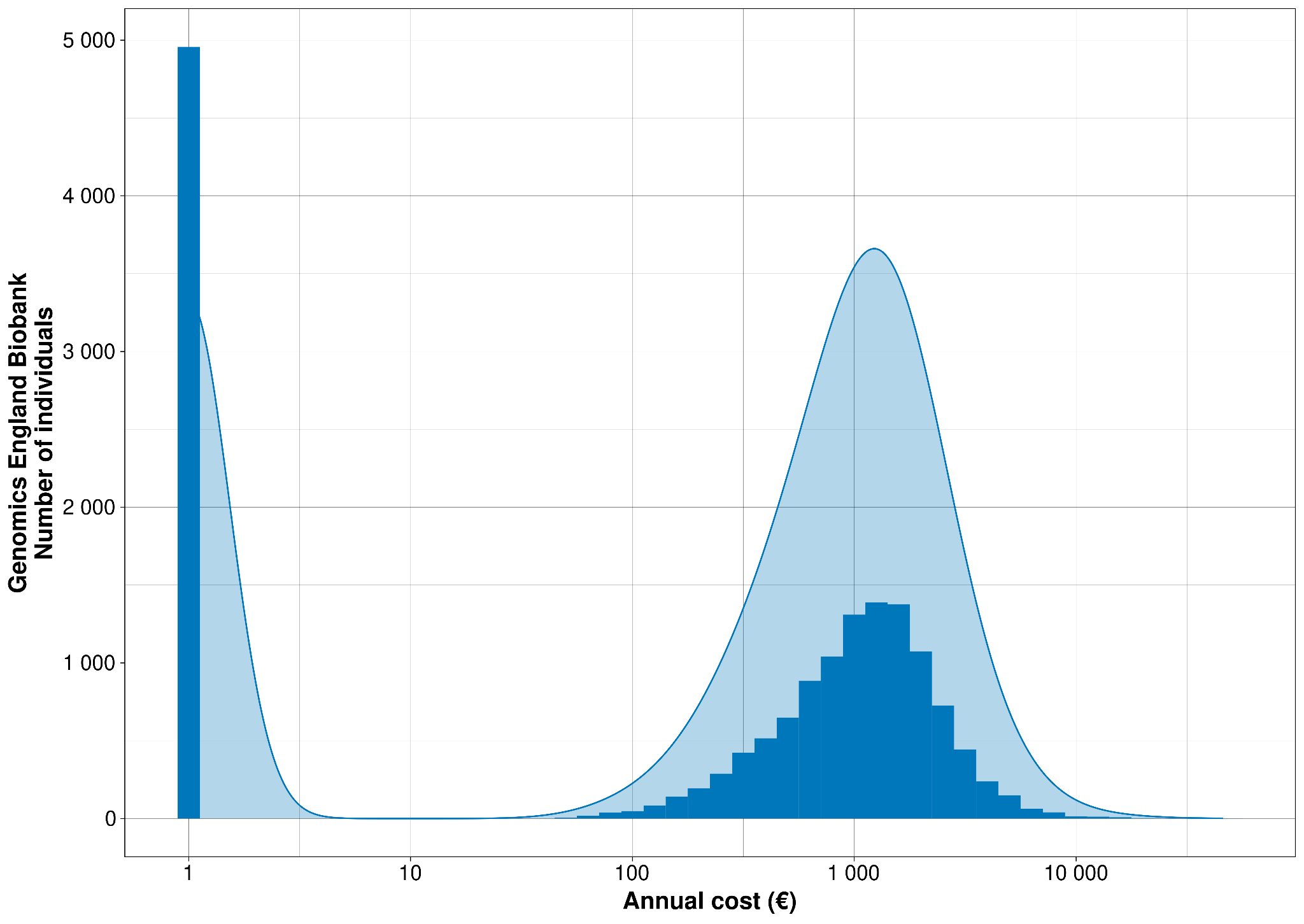

**G - Genes and Health (G&H)**

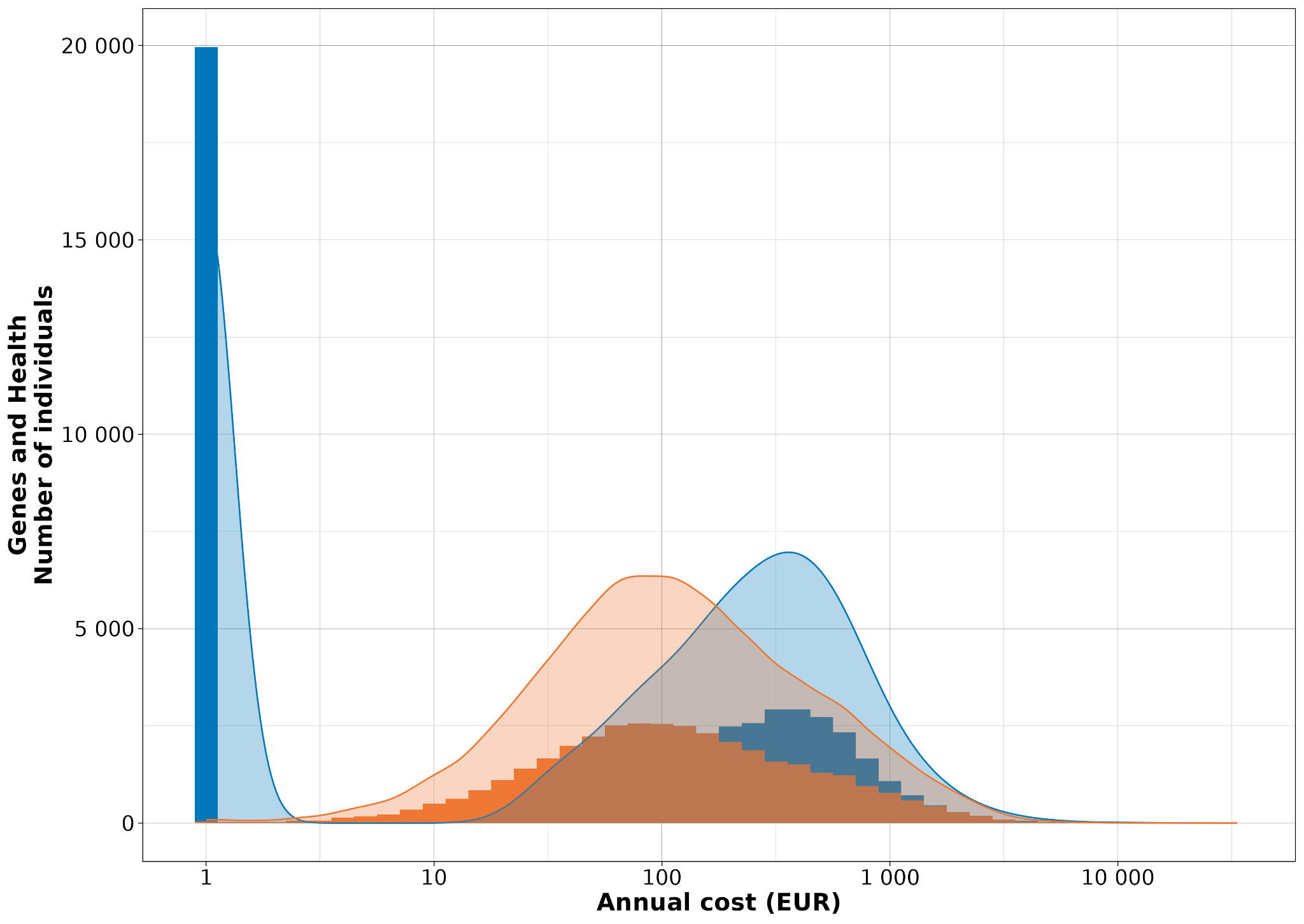

**H - Mass General Brigham Biobank (MGBB)**

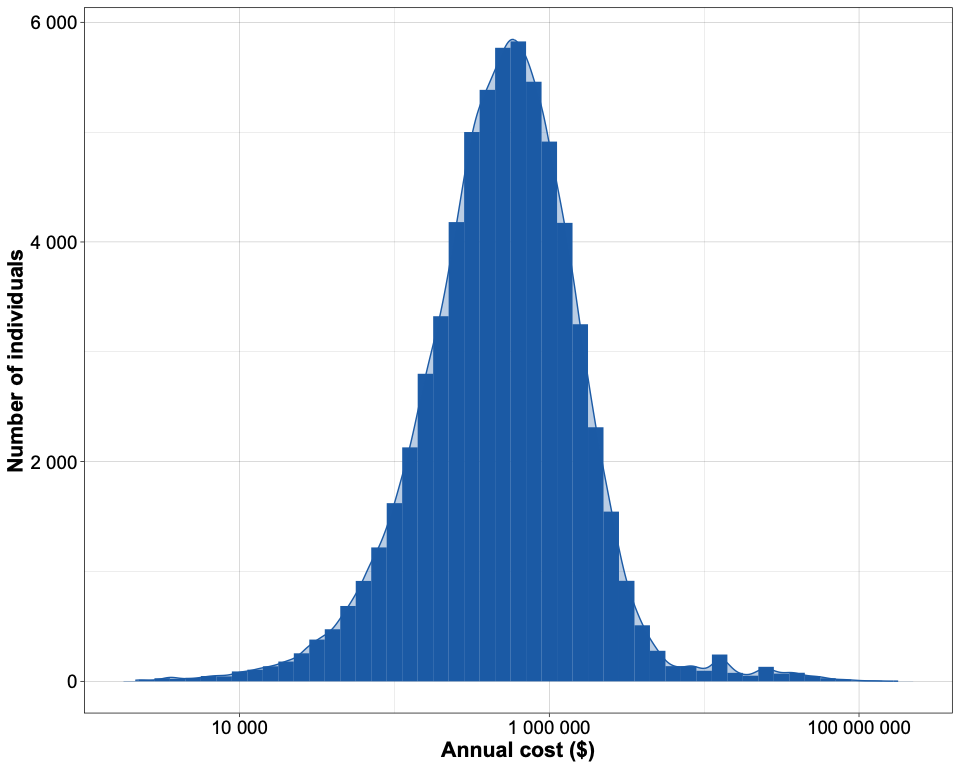

**I - Qatar Genome Project (QGP)**

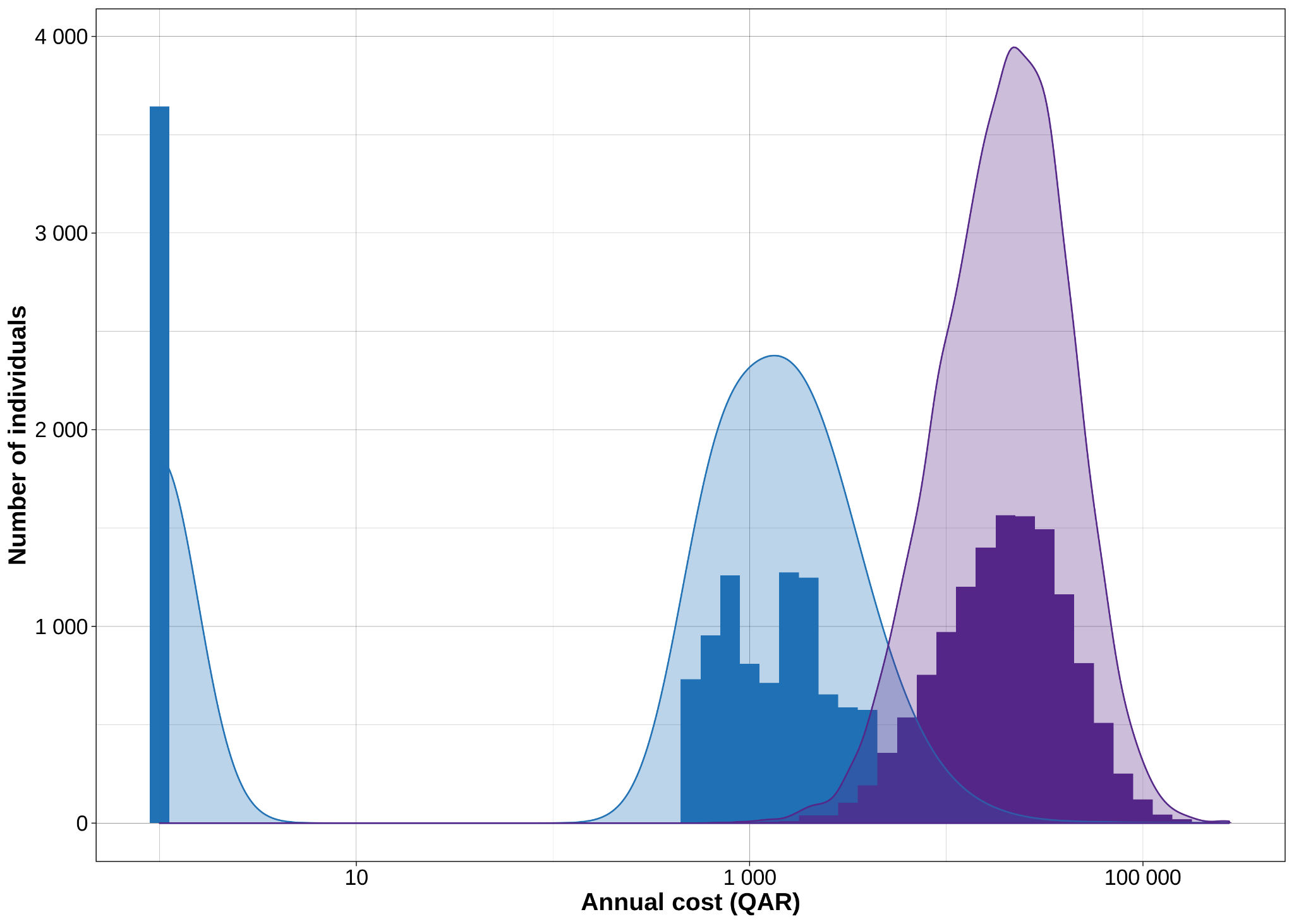

**Supplementary Figure 2.** Miami plot for inpatient + outpatient costs and primary care costs.

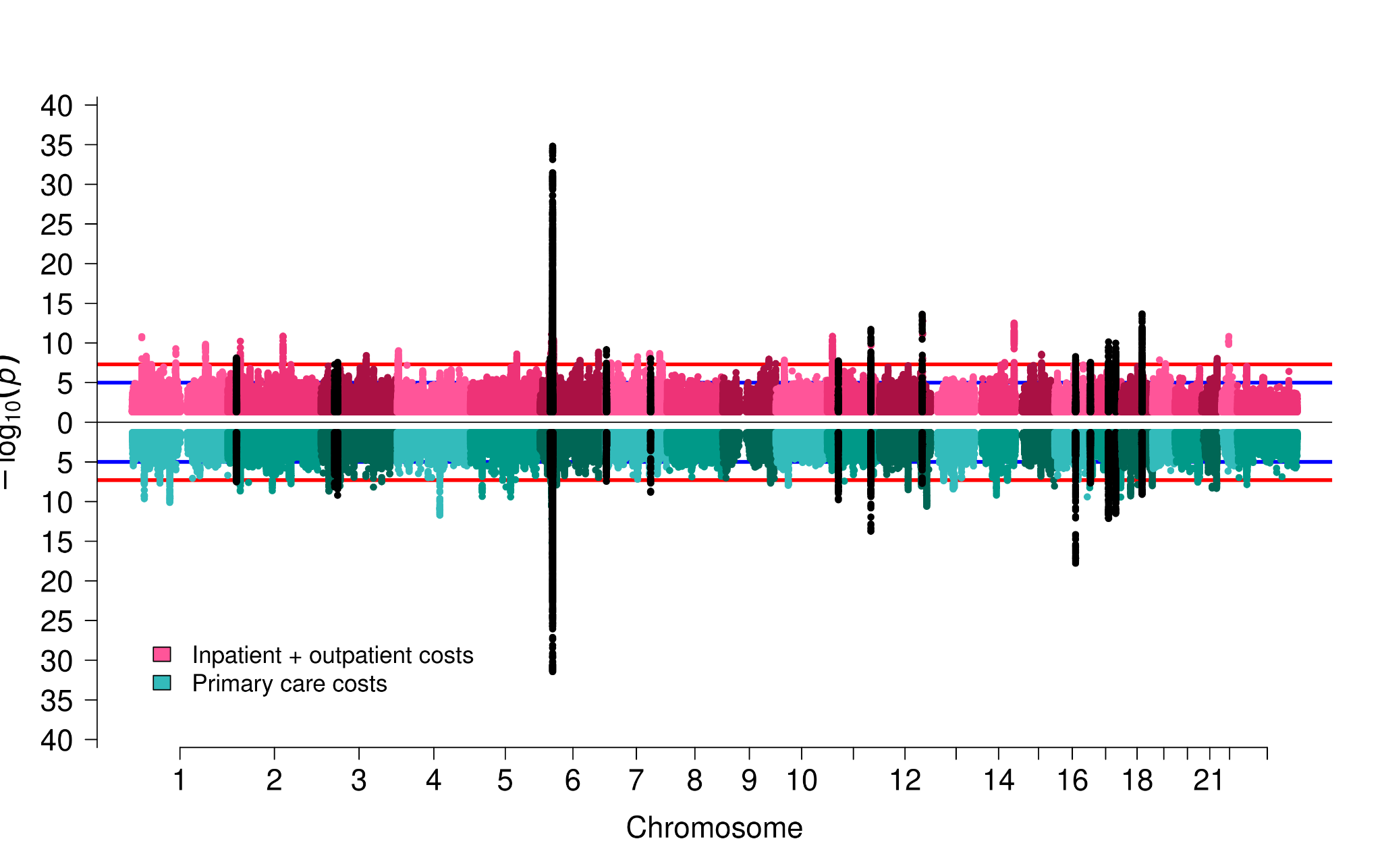

**Supplementary Figure 3.** Upset plot showing overlap of conditionally independent loci across different cost phenotypes.

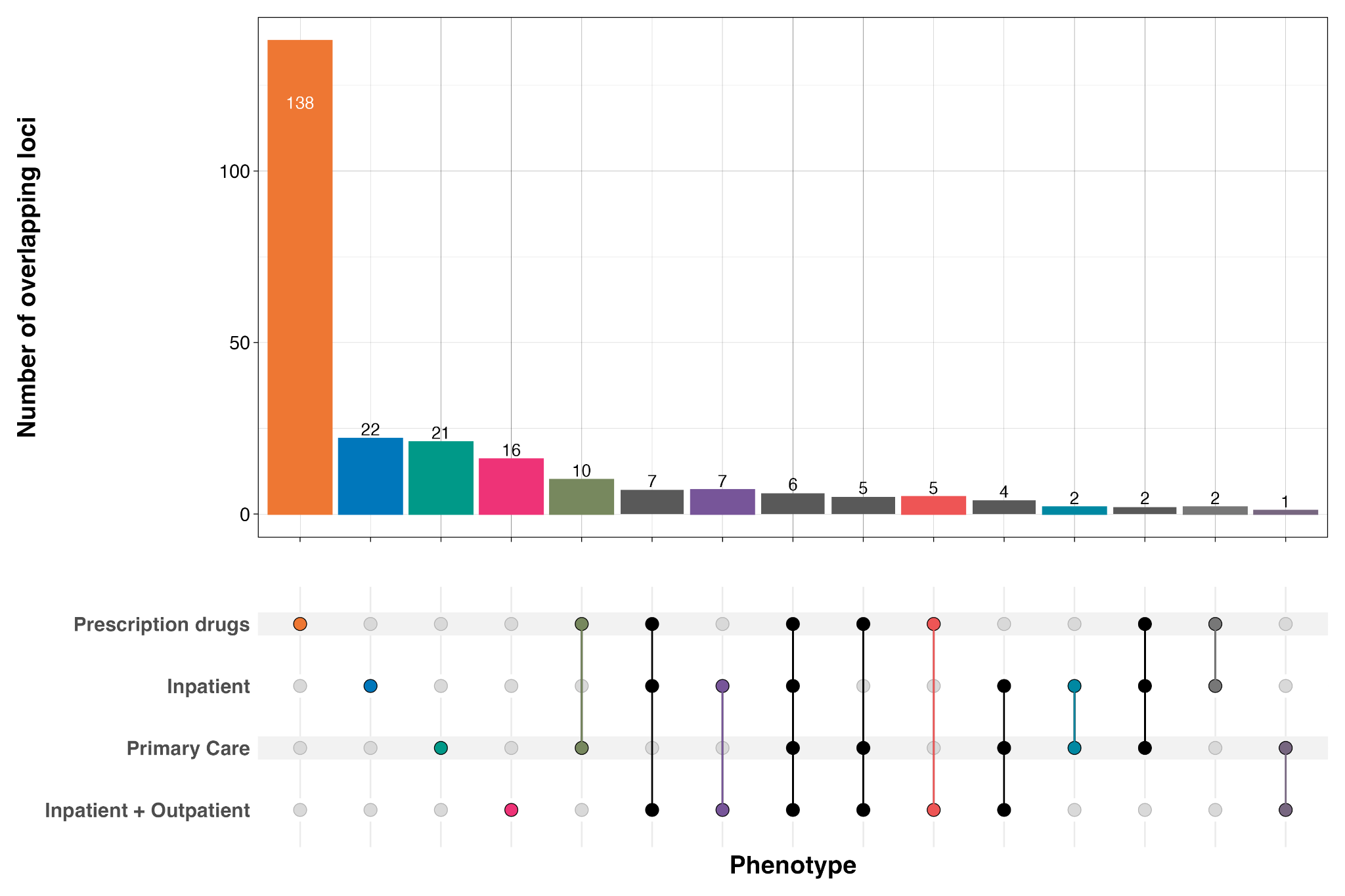
