## Supplementary material for "Quantifying the contribution of genetic variation to healthcare expenditure across diverse healthcare systems": GenCost FAQ

*NOTE: This document is an FAQ to accompany the GenCOST GWAS consortium paper. Here we attempt to anticipate and address potential questions to avoid misunderstandings in the interpretation and use of these results.*

**Why are we studying the genetic basis of healthcare costs?**

Broadly speaking, genetic analyses at a population level have been restricted to specific traits (such as height or BMI) or diseases (e.g. cardiovascular disease, cancer), which is useful for understanding the underlying biology. An individual’s healthcare cost is at its most basic a marker of overall health, with increased expenditures (by the system) in hospital visits or prescription medication potentially indicating worse health and increased risk of morbidity and mortality.

Understanding how much healthcare spending is linked to genetic risk helps researchers build more accurate models of future healthcare costs. It also helps determine when genetic information might actually improve health outcomes. For example, if certain inherited genetic risks are associated with higher medical costs, this information could help decide whether screening programs are worth the investment. While tools like polygenic risk scores (which combine many small genetic effects to estimate disease risk) have generated strong interest, we still do not know how much real-world benefit they provide when used in a clinical setting. Many studies suggesting they are cost-effective rely on simulations or focus on single diseases rather than overall healthcare spending.

Long-term healthcare cost data reflect a person’s overall health journey — including chronic illnesses, sudden medical events, lifestyle habits, and social influences. By studying links between genetics and total healthcare costs over time, researchers may uncover previously unknown or unappreciated biological or behavioural pathways that lead to ongoing illness or heavy use of healthcare services.

**What other similar research has been reported?**

Genetic analysis of healthcare costs has received relatively little attention. We report the largest GWAS of healthcare costs, but two twin studies (one of which included a small GWAS) have also been published. Twin studies rely on the assumption that differences in trait similarity between monozygotic (identical) and dizygotic (fraternal) twins are primarily due to genetic factors. Monozygotic twins share nearly all their genetic variants, while dizygotic twins, on average, share about 50%. Under this assumption — and assuming that both types of twins experience equally similar environments — the proportion of variance in a trait attributable to genetic, shared environmental, and non-shared environmental influences can be calculated.

Lakhani et al (<https://doi.org/10.1038/s41588-018-0313-7>) used an American health insurance dataset to examine the heritability of monthly healthcare cost amongst 56,396 twin pairs. They estimated that the heritability of average monthly cost was 0.290. de Zeeuw et al (<https://pubmed.ncbi.nlm.nih.gov/34213412/>) studied 16,726 participants in the Netherlands Twin Register and estimated a family-level heritability ranging from 0.294 to 0.375. In their small GWAS of 14,572 individuals, no genome-wide significant hits were identified, and SNP (single nucleotide polymorphism) heritability was estimated at between 0.0 and 0.054.

### **Why have you combined costs from different countries with very different health systems?**

Healthcare costs are specific to local contexts. Genetic influences on these costs will be indirect and are likely to be influenced by local healthcare systems including the costs and types of intervention available, local policies, local environments, and cultural norms. Nevertheless, we consider that there is significant value in combining costs from different systems.

In the first instance, larger cross-country samples offer greater statistical power to detect whether any identified genetic signals are likely to be meaningful. A larger sample size also helps with detecting rare variants that may not be identifiable in smaller or single-country cohorts.

Including data from different countries also enhances the ancestral diversity of our cohort. This permits genetic signals from less often studied populations to be included in an international multi-ethnic cohort.

Finally, while health systems and therefore cost phenotypes for the same condition may differ between countries, it is still plausible that the underlying biological and genetic structures influencing morbidity—and therefore healthcare cost—are still the same. This offers some reassurance that any effect isn’t specific to a particular national or health-system context but instead is observable across different institutional environments. It also means our results are more likely to be meaningful for many countries rather than one or two.

### **Why are some SNPs associated with healthcare cost? Can we target these SNPs to make people healthier?**

Much more research is needed to establish why particular SNPs are associated with our outcome. Identified SNPs reflect regions of the genome that influence these costs through many complex biological and social processes that reflect morbidity, hospitalisation, and ultimately treatment costs. These effects may comprise both direct effects, effects of other variants in local linkage disequilibrium with tagged SNPs, and indirect effects that reflect parental or familial behaviour. Although we tried to address some of this with exome sequencing and within-family analysis, much work remains. This work represents an initial start in understanding some of the biology of healthcare costs.

### **Can you predict my healthcare costs using a healthcare cost polygenic risk score?**

No. The genetic variation we report in this paper only explains a small amount (between approximately 0.5 to 2.5%) of overall variation in healthcare costs . Healthcare costs depend on which health system you live in, your health, available treatments, and the costs of preventing and treating ill health. Even then, there can be no guarantee that a prediction for a single individual will be accurate in any practical sense.

### **Can a polygenic risk score calculated for a newborn baby be used to predict their healthcare costs in middle and old age?**

No. On average, other factors will have a much more important influence than an individual's genetic makeup. Genetic variation influences morbidity, but healthcare costs depend in an important way on the price and availability of preventative and therapeutic interventions.

Of course, in some cases—where an individual has a rare genetic variant with extremely severe phenotypic consequences (such as a progressive, lifelong disability, or a massively increased risk of cancer diagnosis)—future healthcare costs may be more predictable. However, genetic information about liability to healthcare costs would not be needed to predict these costs once a diagnosis is established.

Moreover, the future profile of cost will change as new treatments are discovered and made available to the population. This could, for example, plausibly increase costs in some cases, while making the population healthier.

**Can the polygenic risk score indicating higher costs lead to discrimination? What are the implications of our findings for insurance?**

Like many research findings, the results of our study could be misunderstood. As described above, these data cannot and should not be used to predict individual-level healthcare costs. Our results relate to population-level associations and do not readily lend themselves to inferences at the level of the individual.

Nevertheless, the results may raise concerns about their use in insurance, and the potential for individuals with apparently adverse genetic profiles for healthcare cost to be denied insurance, or to have access only to more expensive forms of insurance (https://pubmed.ncbi.nlm.nih.gov/38752549/). On the other side of the market, insurers’ commercial viability may be threatened if consumers—but not providers—can access genetic information (see for example<https://doi.org/10.1101/2025.01.20.25320832>).

Many jurisdictions prohibit the use of genetic information in insurance. However, there are significant international differences in how genetic information can be used. For example, the United Kingdom has a “code” (<https://www.gov.uk/government/publications/code-on-genetic-testing-and-insurance>), agreed between the Government and the representative body for the insurance industry, that prohibits the use of genetic information in insurance for some but not all types of insurance. This code does not apply to all insurers, is primarily focused on underwriting, and does not cover all types of insurance.

In the United States, the Genetic Information Nondiscrimination Act (GINA) was signed into law in 2008. GINA prohibits discrimination based on genetic information in employment and in health insurance. Forms of insurance not covered by the provisions of GINA include life, long-term care, and disability insurance.

The Genetic Non-Discrimination Act in Canada prohibits requesting disclosure of the results of genetic tests or being forced to take such tests in order to obtain access to goods and services including insurance. The Australian life insurance industry introduced a partial, self-regulated ban on the use of genetic results in 2019, and in 2024 the Australian government announced its intention to legislate for such a ban. In November 2025, the Treasury Laws Amendment (Genetic Testing Protections in Life Insurance and Other Measures) Bill 2025 was introduced to the Australian Parliament.

The regulatory landscape across Europe and the European Union is complex. The Oviedo Convention, adopted by the Council of Europe, provides a framework for the use of genetic data in healthcare and research. Although not ratified by all European countries, it prohibits “discrimination” based on genetics. In March 2025, the European Union adopted the European Health Data Space (EHDS) Regulation. The Regulation defines “health data” broadly to include electronic health data such as human genetic, genomic, and epigenomic data. Article 54 addresses the secondary use of health data accessed through the EHDS framework and prohibits insurers (and other secondary users) from offering less favourable terms or excluding individuals or groups from contracts — including insurance and credit — on the basis of such health data.... "

**I am a participant in one of the studies or biobanks that you included in your study. Can my insurers now access my personal cost data?**

No, for several reasons. Firstly, the results of the GenCost project relies on aggregated summary-level data, which does not incorporate data about any individual, making it impossible to identify any individual whose healthcare costs were used in the project.

Access to the underlying data used in this GWAS is governed by the policies of each contributing study, and such access is strictly regulated. Identifying individual participants is explicitly restricted by access committees and is only allowed under exceptional circumstances. Insurers may be able to use study data for population-level research, but this is entirely distinct from using such data to determine pricing or contract terms for any specific individual—even if that individual’s data were included in the study.

In most cases, the cost phenotype that we analyse as our primary outcome measure was specifically developed for the purposes of this study and does not preexist in the original datasets from which we drew data. In any event, any applicant seeking to use data from these studies would still need to apply for access through the proper channels.
