## Supplementary material for "Quantifying the contribution of genetic variation to healthcare expenditure across diverse healthcare systems": Banner Authors

**Extended cohort authors**

Additional banner authors and contributors to the studies involved in GenCost are detailed here.

**FinnGen**

See attached FinnGen banner author table.

**Estonian Biobank**

**Banner Authors**

Andres Metspalu¹, Mait Metspalu¹, Lili Milani¹, Reedik Mägi¹, Mari Nelis¹, Georgi Hudjashov¹, Tõnu Esko¹

¹Estonian Genome Center, Institute of Genomics, University of Tartu, Tartu, Estonia

**Genes and Health**

**Banner Authors**

Eamonn Maher¹, Shabana Chaudhary², Joseph Gafton², Karen A. Hunt², Shapna Hussain², Kamrul Islam², Mohammed Bodrul Mazid², Elizabeth Owor², Jessry Russell², Nishat Safa², John Solly², Marie Spreckley², David A. Van Heel², Jan Whalley², Ishevanhu Zengeya², Emily Mantle², Shaheen Akhtar³, Samina Ashraf³, Dan Mason³, John Wright³, Daniel MacArthur⁴, Michael Simpson⁵, Richard C. Trembath⁵, Gerome Breen⁵, Raymond Chung⁵, Sang Hyuck Lee⁵, Omar Asgar⁶, Joanne Harvey⁶, Karen Tricker⁶, Caroline Winckley⁶, Hanifa Khatun⁶, Amna Asif⁶, Claudia Langenberg⁷, Grainne Colligan⁸, Ceri Durham⁸, Bill Newman⁹, Ahsan Khan¹⁰, Hilary Martin¹¹, Teng Heng¹¹, Matt Hurles¹¹, Vivek Iyer¹¹, Georgios Kalantzis¹¹, Vladimir Ovchinnikov¹¹, Iaroslav Popov¹¹, Klaudia Walter¹¹, Panos Deloukas¹², David Collier¹², Ana Angel¹³, Saeed Bidi¹³, Fabiola Eto¹³, Sarah Finer¹³, Chris Griffiths¹³, Sam Hodgson¹³, Benjamin M. Jacobs¹³, Rohini Mathur¹³, Caroline Morton¹³, Asma Qureshi¹³, Stuart Rison¹³, Annum Salman¹³, Miriam Samuel¹³, Moneeza K. Siddiqui¹³, Daniel Stow¹³, Sabina Yasmin¹³, Julia Zöllner¹³, Sheik Dowlut¹³

¹Aston University
 ²Blizard Institute, Queen Mary University of London
 ³Bradford Teaching Hospitals NHS Foundation Trust
 ⁴Garvan Institute
 ⁵King’s College London
 ⁶Manchester University Hospitals
 ⁷Precision Healthcare University Research Institute, Queen Mary University of London
 ⁸Social Action for Health (charity)
 ⁹University of Manchester
 ¹⁰Waltham Forest Council
 ¹¹Wellcome Sanger Institute
 ¹²William Harvey Research Institute, Queen Mary University of London
 ¹³Wolfson Institute of Population Health, Queen Mary University of London

**China Kadoorie Biobank**

**Members of the China Kadoorie Biobank collaborative group**

**Steering Committee:** Junshi Chen, Zhengming Chen (PI), Rory Collins, Liming Li (PI), Jun Lv, Richard Peto, Robin Walters.

**National Co-ordinating Centre, Beijing:** Liming Li, Jun Lv, Canqing Yu, Dianjianyi Sun, Yuanjie Pang, Yuting Han, Can Hou, Qingmei Xia, Chao Liu, Pei Pei, Lang Pan, Xiao Han, Honglu Bian, Xinxin Chen.

**International Co-ordinating Centre, Oxford:** Daniel Avery, Maxim Barnard, Derrick Bennett, Ruth Boxall, Yiping Chen, Zhengming Chen, Jonathan Clarke, Huaidong Du, Ahmed Edris, Hannah Fry, Pek Kei Im, Andri Iona, Christiana Kartsonaki, Kshitij Kolhe, Hubert Lam, Kuang Lin, James Liu, Iona Millwood, Sam Morris, Qunhua Nie, Alfred Pozarickij, Maryam Rahmati, Paul Ryder, Maruf Sarder, Dan Schmidt, Becky Stevens, Robin Walters, Baihan Wang, Lin Wang, Neil Wright, Ling Yang, Xiaoming Yang, Pang Yao.

**Regional Co-ordinating Centres:** **Qingdao CDC:** Zengchang Pang, Ruqin Gao, Kunzheng Lv, Shanpeng Li, Haiping Duan, Shaojie Wang, Yongmei Liu, Ranran Du, Liang Cheng, Xiaocao Tian, Hua Zhang. **Licang CDC:** Dan Hu, Xiaoyan Zheng, Yujie Wang. **Heilongjiang Provincial CDC:** Wei Sun, Shichun Yan, Yong Zhou. **Nangang CDC:** Chi Wang, Zhenyuan Wu, Lishun Zhai, Zhaoxi Pang, Shiwen Dong, Li Liu. **Hainan Provincial CDC:** Dapeng Yin, Bin He, Ying Liu, Xingren Wang, Tingting Ou. **Meilan CDC:** Xiangyang Zheng, Dewei Zheng, Shuai Yang, Lihui Li, Xingjiao Chen. **Jiangsu Provincial CDC:** Yan Xu, Jinyi Zhou, Ran Tao, Jian Su, Xikang Fan, Xuejia Chen, Yuxiao Huang. **Suzhou CDC:** Yan Lu, Yujie Hua, Li Xing, Shuxian Wang, Jianrong Jin, Juping Ma, Jinchao Liu, Kaifei Zhu, Hongfu Ren, Xingfeng Shen. Guangxi **Provincial CDC:** Ge Zhong, Wei Mao, Zhenzhen Lu, Yanxu Zhong. **Liuzhou CDC:** Lifang Zhou, Rong Pan, Jian Lan, Xiaoping Tan, Jinxue Tan, Yishan Xie, Liuping Wei, Liyuan Zhou, Sisi Wang. **Sichuan Provincial CDC:** Xianping Wu, Ningmei Zhang, Xiaofang Chen, Xiaoyu Chang, Zhuo Wang, Yujin He. **Pengzhou CDC:** Mingqiang Yuan, Ling Wang, Xiaofang Chen, Zhaodong Wang, Qiang Sun, Yang Lin. **Gansu Provincial CDC:** Faqing Chen, Xiaolan Ren, Lijun Chang, Feiming Zhong. **Maiji CDC:** Jianjun Feng, Weijie Hu, Xiaofang Zhang, Yalin Chen, Honghong Wang, Jun Wang. **Henan Provincial CDC:** Linqi Diao, Zhiwei Han, Dengjun Zhu, Kai Kang, Shixian Feng, Wenjie Yang, Huizi Tian, Yali Yan, Bing Han, Li Gao, Shaofang Li, Tianfang Xing, Wei Tang. **Huixian CDC:** Xiaolin Li, Huarong Sun, Xiaocong Zhao, Ying Li, Chen Hu, Pan He, Xukui Zhang, Yuanyuan Jin, Lan Luo. **Zhejiang Provincial CDC:** Min Yu, Ruying Hu, Hao Wang, Weiwei Gong, Jieming Zhong, Meng Wang, Chunxiao Xu, Keqing Gong. **Tongxiang CDC:** Hao Xu, Yuan Cao, Kaixu Xie, Lingli Chen, Xiaomei Tu, Junlong Pan. **Hunan Provincial CDC:** Xiaojun Li, Li Yin, Huilin Liu, Yuan Liu, Lei Yin, Xian Xie, Jing Wang. **Liuyang CDC:** Bo Xiao, Zongwei Deng, Yuan Peng, Libo Zhang, Chan Qu, li Deng, Qili Jiang, Yanling Chen.
